# A Systematic Review of Human Studies Assessing the Health Effects of Kerosene-Based Jet Fuels and Products Across Diverse Populations and Settings

**DOI:** 10.1101/2025.08.12.25333531

**Authors:** V. Carvajal, B. Ng, N. Rosendaal, C. M. Pirkle

## Abstract

**Objective:** Total petroleum hydrocarbons are a significant environmental health concern globally, due to their extensive consumption and environmental release. However, their toxicological mechanisms and associated health consequences, particularly in their unburned forms, are poorly understood in humans. A notable gap is research on kerosene and kerosene-based jet fuels, with the latter accounting for 8% of total daily petroleum consumption.While previous reviews have examined health effects associated with these fuels, the focus on occupational settings and post-combustion forms limit our understanding of potential health implications. Recent drinking water contamination events which expand beyond the scope of existing research highlight these limitations, particularly for other settings and underrepresented groups. Therefore, we conducted a systematic review of the evidence of human health impacts of pre-combustion forms of kerosene and kerosene-based jet fuels across all exposure settings and population groups.

**Material and Methods:** With the assistance of a public health librarian, a search strategy, search terms, and eligibility criteria were developed and executed following PRISMA guidelines. PubMed and Web of Science databases were used to search for eligible literature published between 2017 and 2024. Data extraction, analyses and quality assessment were conducted. Three reviewers participated in this review.

**Results:** 28 articles were included. Limited evidence of a casual relationship between unburned kerosene and kerosene-based jet fuel and health outcomes were identified. Current research suggests that these fuels affect respiratory and neurological systems, but there was also evidence for gastrointestinal and dermatological outcomes. Respiratory effects were frequently observed following acute kerosene ingestion, whereas neurological health outcomes were common with chronic occupational jet fuel exposure, particularly through dermal and inhalation routes. The limited number of analytical studies available, and lack of consistency in exposure and outcome variables, challenge meaningful conclusions.

**Conclusion:** The evidence identified by this review was limited. We provide recommendations for future studies, covering aspects such as exposure assessment methods, study designs, exposure context, and prioritization of underrepresented populations. Considering the global health implications of kerosene and kerosene-based jet fuels, and recent contamination events that underscore the limitations of existing research, furthering our understanding of human health risks associated with these fuels should be prioritized.

## 1. Introduction

### 1.1. Total Petroleum Hydrocarbons as Global Environmental Health Concern

Petroleum hydrocarbons (PH) - the family comprising hundreds of crude oil derived chemical compounds - are frequently described as the most prevalent environmental contaminants globally (Parmar et al., 2024; Truskewycz et al., 2019; Zhang et al., 2013), due to their extensive anthropocentric uses in transportation, heating, and industry (Agency for Toxic Substances and Disease Registry (ATSDR), 1999; Kuppusamy et al., 2020). Global petroleum consumption reached 97.3 million barrels per day (b/d) in 2021 (U.S. Energy Information Administration (EIA), 2023). Total petroleum hydrocarbons (TPH) refer to the combined measurable concentration of PHs in a sample, and varies with the analytical method used, environmental medium, and extractable components of the contaminating petroleum mixture (ATSDR, 1999; Kuppusamy et al., 2020). Through the refinement of crude oil, operational failures, leakages, spills, and as byproducts from commercial or private uses, TPHs often enter the environment, contaminating water, soil and air (ATSDR, 1999; Kuppusamy et al., 2020). When released into water, lighter TPH fractions may float, while heavier fractions accumulate in sediment, potentially impacting wildlife who inhabit affected areas, and those that feed on them, resulting in bioaccumulation (ATSDR, 1999; Kuppusamy et al., 2020) and potential human health implications following consumption of these organisms. In soil, TPHs can seep from the source area, evaporate into air, and/or dissolve into groundwater (ATSDR, 1999). In other instances, TPH compounds may enter groundwater directly (Cavanaugh, 2022).

Considering that TPHs commonly spread from the area in which they are released, people may come into contact with these compounds even when at distance from the pollution site (Kuppusamy et al., 2020). TPHs may enter the body when inhaled, ingested, or in contact with the skin (ATSDR, 1999), and impact various organ systems depending on how they are metabolized and distributed (ATSDR, 1999). The resulting health effects are dependent on the nature of the compounds themselves, the duration of exposure, as well as the exposure dose (ATSDR, 1999). While the toxicity of particular fractions such as benzene, toluene, ethylbenzene and xylenes are well described, the human health effects associated with exposure to more complex fuel mixtures (ATSDR, 1999; Kuppusamy et al., 2020), such as jet fuel and kerosene, are less understood despite their extensive use and frequent release into the environment.

### 1.2. Kerosene & Jet Fuel: Composition, Use and Limitations of Existing Research

In 2022, daily petroleum consumption averaged approximately 20.3 million b/d in the United States alone, for industry, residential and commercial purposes (U.S. EIA, 2023). More than half of this use, (67%) went to transportation including the 1.6 million barrels of kerosene-type jet fuel that accounted for 8% of total daily petroleum consumption (U.S. EIA, 2023). In contrast, kerosene as a standalone product was consumed at a much lower rate of 0.004 million b/d (U.S. EIA, 2023).

Kerosene (commonly known as fuel oil no. 1, paraffin, paraffin oil, lamp oil) is a colourless to yellow appearing petroleum-derived oil used as household fuel for heaters, lamps, furnaces, diesel and tractor engines, as well as in lubricants and pesticides (ATSDR, 1999; Kuppusamy et al., 2020; Lam et al., 2012; National Institute for Occupational Safety and Health (NIOSH), 2019). In lower-and-middle-income countries where access to electricity is not always available, kerosene is often used as fuel for cooking and lighting, contributing to air pollution that has been associated with several adverse health outcomes (Lam et al., 2012; Puzzolo et al., 2024). Thus, studies examining the health effects of kerosene use in the home generally assess outcomes related to combustion-specific byproducts, without consideration of potential risks of exposure to raw, unburned fuel. Adverse human health effects of exposure to kerosene pre-combustion are documented; however, the available evidence is limited and stems primarily from reports of accidental ingestion in children of low- and middle-income countries (ATSDR, 1999; Ritchie et al., 2011)

While residential usage has steadily declined in higher income countries, globally, kerosene continues to be used frequently in commercial and military aviation, predominantly as jet fuel (Kuppusamy et al., 2020; Lam et al., 2012). Although a small amount of jet fuel is produced from oil sands, the vast majority is kerosene based (ATSDR, 2017; Chevron, 2006). These kerosene-type jet fuels contain varying proportions of kerosene itself, along with performance additives, including antioxidants and agents to prevent icing, static buildup, corrosion and bacterial growth (ATSDR, 1999; Kuppusamy et al., 2020). Two major kerosene-based jet fuels are used in commercial aviation: Jet A and Jet A-1. Jet A-1 has a lower maximum freezing point suitable for international flights. Jet A is used commercially for flights within the continental United States and by the U.S. Air Force (ATSDR, 2017). Military jet propellants (JP) include JP-4, 5, 6, 7, 8 (ATSDR, 2017), each differing in intended application and chemical composition (Kuppusamy et al., 2020). Of these, JP-5 and JP-8 are the two kerosene-based aircraft fuels used currently by the U.S. military and contain approximately 99.5% kerosene (ATSDR, 2017; Kuppusamy et al., 2020). Due to the increased safety associated with its higher flash point (i.e. the lowest temperature at which a liquid can be ignited when an ignition source is present), JP-5 is used by the U.S. Navy aboard aircraft carriers (ATSDR, 2017). JP-8 is chemically similar to Jet A-1, but with military-specific performance additives, and is the most widely used jet fuel by the U.S. military in both air and ground applications, including fuel for aircraft, automotives, heaters, lighting and other equipment (ATSDR, 2017; Ritchie et al., 2003).

Jet fuels (including Jet A, JP-5 and JP-8) often enter the environment during routine processes such as storage, handling and transportation, in-flight jettisoning, as well as accidental spills or leaks (ATSDR, 2017). Like other TPHs, jet fuel can migrate from the pollution site, with the potential to penetrate soil, enter groundwater, and/or be carried by the atmosphere following volatilization (ATSDR, 2017).

Jet fuel is the largest chemical exposure among U.S. military personnel, and a frequent occupational exposure in non-military settings, particularly for those involved in fueling, transporting and routine maintenance of aircraft (ATSDR, 2017; Ritchie et al., 2003). These individuals are at an increased risk, particularly through dermal absorption and inhalation of both pre- and post- combustion fuel forms (ATSDR, 2017). This elevated risk is reflected in current research, where the limited number of human studies assessing jet fuel associated health outcomes are predominantly focused on occupational exposures (ATSDR, 2017; Ritchie et al., 2003; U.S. Department of Veterans Affairs, 2023; Vincent-Hall et al., 2025). It has been recognized in the literature, however, that the general population residing in close proximity to military bases and airports are at increased risk of exposure to jet fuel and toxic effects from them (ATSDR, 2017; Bendtsen et al., 2021; Ritchie et al., 2003).

The toxic effects of jet fuel exposure and the physiological mechanisms behind those effects are understudied (ATSDR, 2017; Ben Maamar et al., 2020; Ritchie et al., 2003). The current body of research on this topic consists largely of animal or *in vitro* studies. Despite these gaps - as well as the frequent and widespread release of jet fuels into the environment, and their established potential to disperse through air, soil and water - the risk of exposure to the general population has been characterized as low (ATSDR, 2017). In 2017, the Agency for Toxic Substances and Disease Registry (ATSDR) published a toxicological profile which consolidated the existing research surrounding potentially-associated health effects of jet fuel. While their scope was not exclusive, the limited human-specific data available were derived primarily from occupational reports. Additional reviews specific to human health outcomes have been published since; however, the focus of these studies were restricted to occupational and/or military settings. As a result, much of the evidence informing our current knowledge of health outcomes associated with jet fuel exposure in humans is based on inhalation and dermal exposures among men of working age (Ritchie et al., 2003; U.S. Department of Veterans Affairs, 2023; Vincent-Hall et al., 2025). There remains a critical gap in understanding the potential health effects of residential, community and oral exposures, particularly in sub-populations including women, children, and older adults. Many of the occupational studies conducted thus far have not distinguished between pre- or post-combustion exposures, ultimately limiting the ability to attribute toxicological outcomes to specific forms of the fuel and their unique chemical compositions.

Understanding health effects specific to raw jet fuel is increasingly urgent in light of recent contamination events. In November 2021, a spill of 5,542 gallons of the kerosene-based JP-5 at the U.S. Navy’s Red Hill Bulk Fuel Storage Facility on Oʻahu, Hawaiʻi contaminated the Navy-managed system supplying drinking water to Joint Base Pearl Harbor Hickam and surrounding communities (Cavanaugh, 2022). Approximately 93,000 individuals, ranging in age from less than a year to over 65 years of age (U.S. Environmental Protection Agency (EPA), 2024; Miko et al., 2023) were exposed to the jet fuel-contaminated water in their households, schools, daycares and workplaces (U.S. EPA, 2024). This event garnered extensive global attention (Alfonsi, 2024; Associated Press, 2021; Cramer, 2021), with the resulting defueling of the facility and ongoing legal battles continuing to be covered extensively by the media (Boone, 2025; Cohen, 2024; Ives, 2022). More recently, in Bucks County, Pennsylvania, a jet fuel leak was confirmed in January 2025, in which JP-8 emanated from Sunoco’s Twin Oaks pipeline and tainted well water (Pipeline and Hazardous Materials Safety Administration, 2025; Wright, 2025). The leak is believed to have been ongoing for at least 16 months (Axelrod, 2025; Pinder, 2025). These events highlight jet fuel exposures in the general population, specifically through ingestion, topics that are both understudied.

### 1.3 Objective

Considering the widespread and frequent release of jet fuel into the environment, the historical focus on males in occupational settings and the limited understanding of health effects associated with pre-combustion jet fuel exposure in humans, a systematic review designed to capture information on these gaps is warranted. Given that kerosene constitutes approximately 99.5% of jet fuel, its inclusion in this review enables examination of this toxicant across a broader range of exposure routes and populations that would remain unassessed with a focus on jet fuel alone. As the literature on jet fuel primarily involves inhalation and dermal contact among occupationally exposed men of working age - with little delineation between pre- versus post-combustion exposures - incorporating kerosene allows for an evaluation of potential health effects linked to ingestion, in additional contexts and demographic groups, to raw, unburned fuel.

Thus, this review aims to systematically evaluate human studies that assess the association between oral, dermal and inhalation exposures to pre-combustion kerosene-based jet fuel, as well as other kerosene products, on health outcomes across all exposure settings and population groups.

## 2. Methods

To guide our search strategy and eligibility criteria, we structured our research question following the PECO framework (Population, Exposure, Comparator, Outcome):*“What are the known and/or potential health effects of exposure (oral, dermal and inhalation) to pre-combustion forms of kerosene-based jet fuel, and other kerosene products, in humans across all exposure settings and population groups?”.* Upon completing our search protocol, the review was registered in The International Prospective Register of Systematic Reviews (PROSPERO) (CRD42025645179). Preferred Reporting Items for Systematic reviews and Meta-Analyses (PRISMA) guidelines were followed in the creation and execution of the methodology for the present review.

### 2.1 Search Strategy and Eligibility Criteria

Databases utilized to conduct literature searches included PubMed and Web of Science. In collaboration with a university public health librarian, the search strategy was designed with the following structure: (Jet fuel & Kerosene terms) AND (Health effects terms). Detailed search strategies (including exposure and health outcome terms) as well as limits (publication years and article types) utilized for each database search can be found in Table 1 (search conducted February 3, 2025). Health effect terms were screened for added value to the search strategy; those terms that did not pull additional results for the search strategy were not included. These terms can be found in Table S1. Additionally, we conducted retrospective citation tracing of included publications.

**Table 1.**
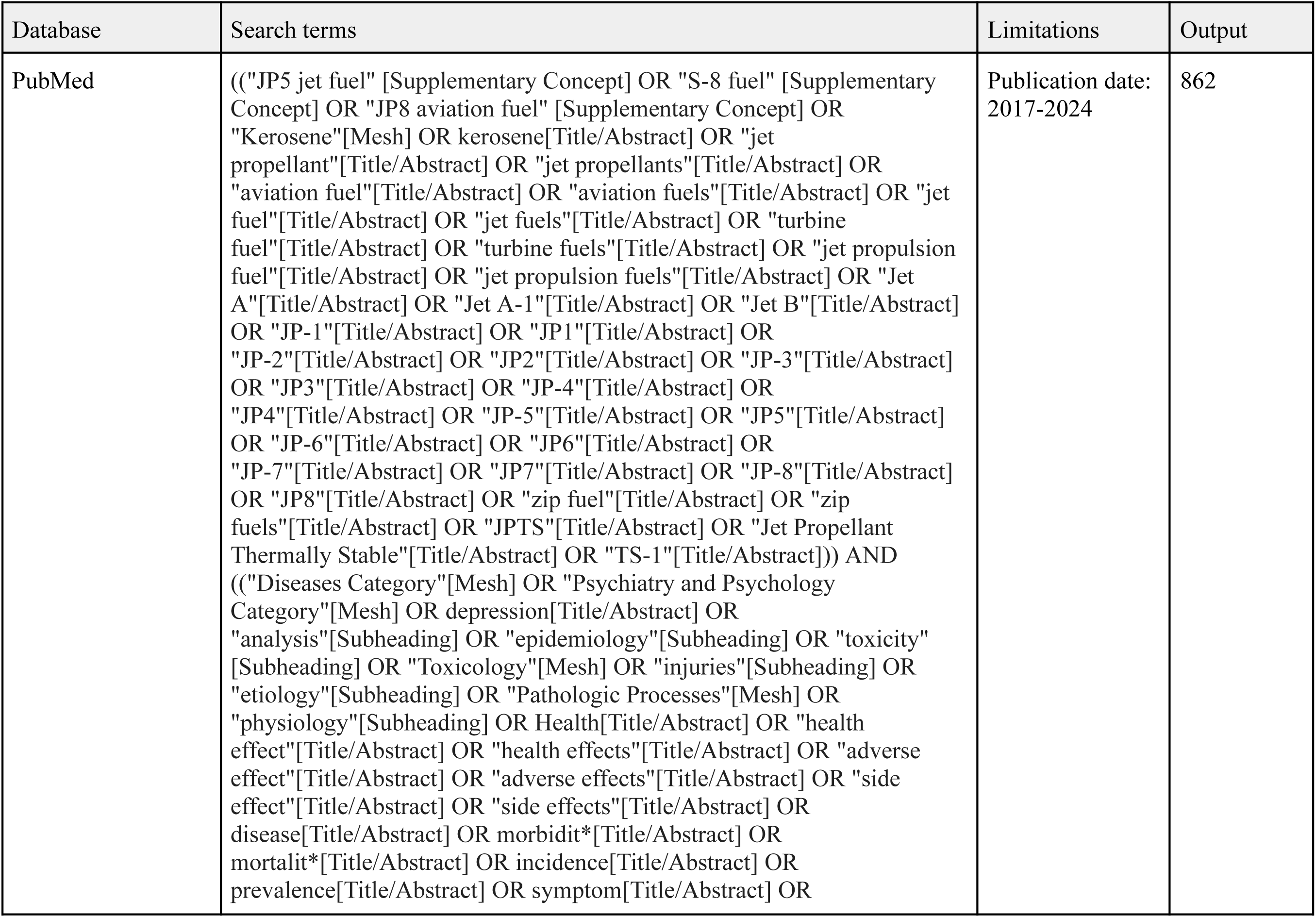
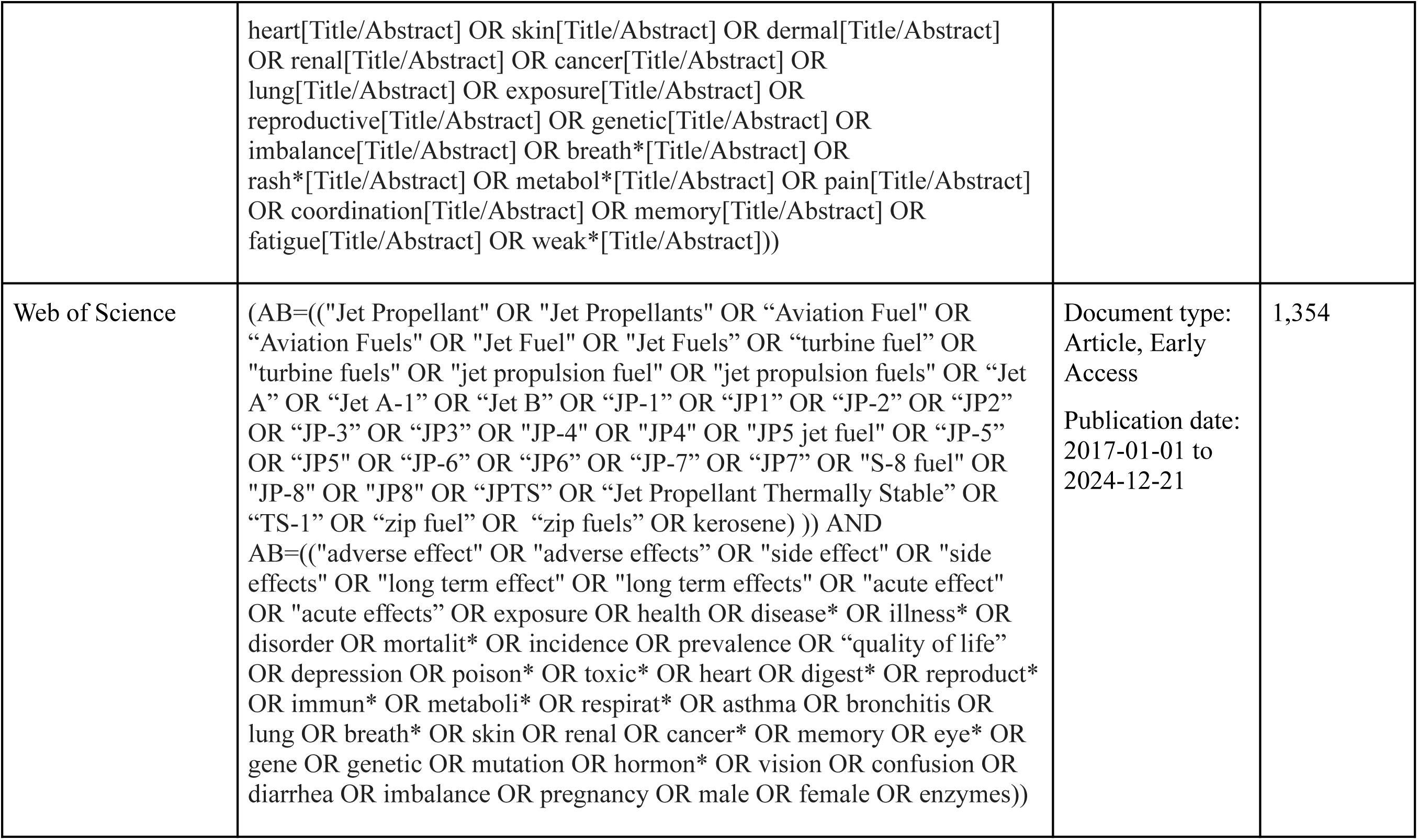
Databases and Search Strategies Used.

For an article to be eligible for inclusion in this review, it had to: a) assess the association between oral, dermal and/or inhalation exposures to pre-combustion forms (i.e. before fuel is burned) of kerosene based jet fuel or other kerosene products, and health related effects in humans; studies focused solely on animal models or post-combustion forms of jet fuel were excluded; b) be peer reviewed and an original study; grey literature, letters to editors, editorials and reviews were excluded; c) be published between 2017 to 2024 in English, French, Dutch, Spanish or Portuguese; d) be experimental, descriptive or observational in design, including but not limited to randomized control trials, case studies as well as case-control and cohort studies; e) conduct investigations *in-vivo*; studies conducted *ex-vivo* were excluded. The publication period was selected to build upon the evidence consolidated by the U.S. ATSDR in their peer-reviewed toxicological profile on jet fuels published in 2017. No limitations were placed on the region in which the studies were conducted. While texts were eligible for inclusion if in French, Dutch, Spanish or Portuguese, searches were conducted in English.

Articles retrieved from the database searches were uploaded to the Rayyan platform for de-duplication, screening and selection of articles. Two reviewers (VC and BN) conducted blind independent screening of all titles and abstracts, as well as full texts. Reviewers met after each stage of the screening process to discuss any conflicting decisions on article eligibility. A third reviewer (CP) was consulted to finalize the decision on eligibility of 2 articles when consensus could not be reached between the first and second reviewers. The article selection process can be found in Figure 1.

**Figure 1.**
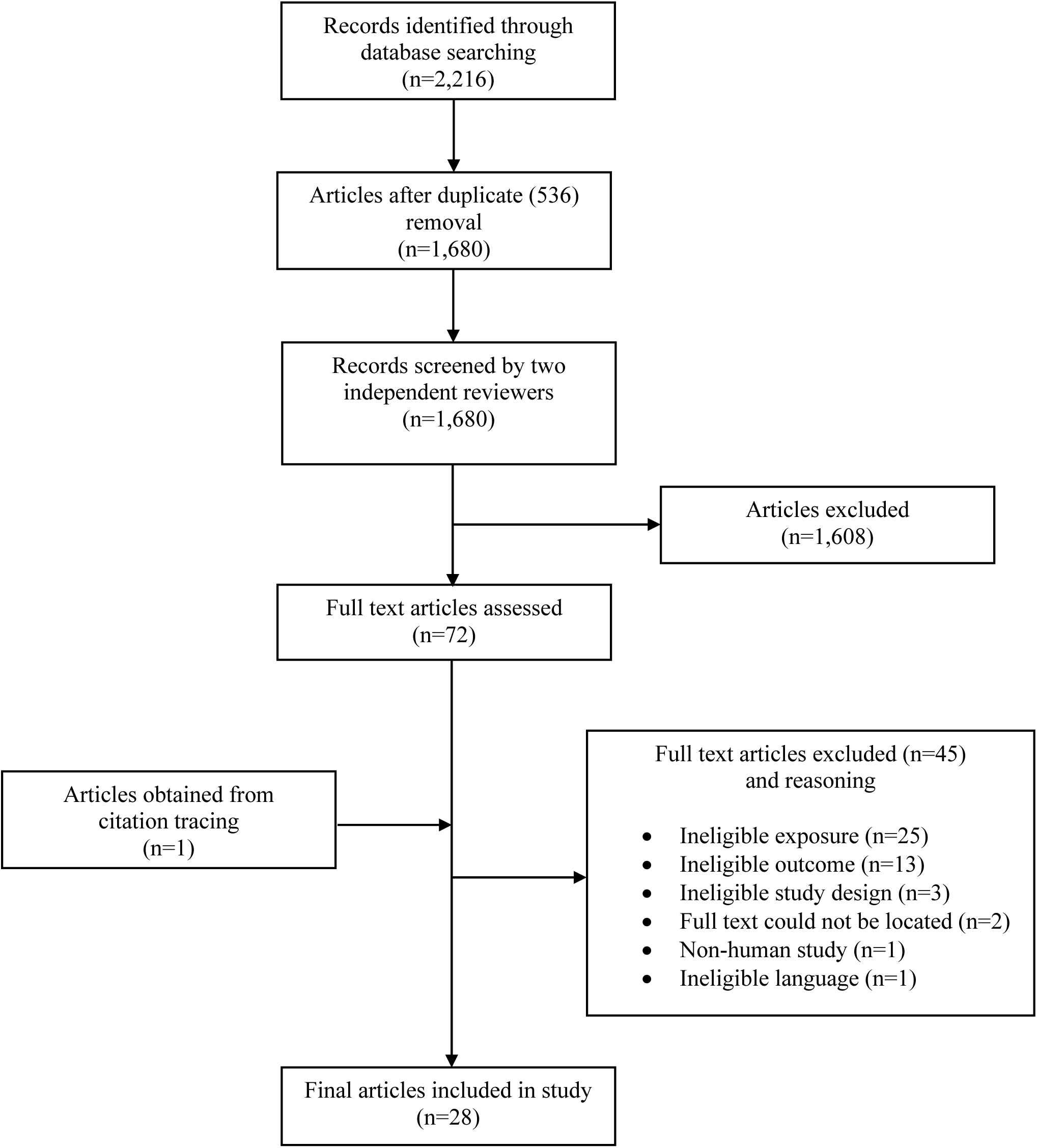
Article Selection Process.

### 2.2 Data Extraction

Data extracted from included articles consisted of: first author, publication year, country in which the study was conducted, study design, sample size, age and sex of participants, type, route and setting of exposure, exposure assessment methods, health outcomes assessed, outcome assessment methods, statistical models, measures of effect, covariates adjusted for in statistical analyses, and main findings applicable to the research question of this review. Study designs were recorded as stated by the original authors. Each included study was data extracted blindly and independently by two reviewers (VC and BN). Any discrepancies in extracted data by reviewers was resolved through discussion to ensure the accuracy and consistency of findings. If data were missing from a given study, it was recorded as “Information not provided” and accounted for when conducting quality assessment of included studies. Extracted data for each study can be found in Table 2.

**Table 2.**
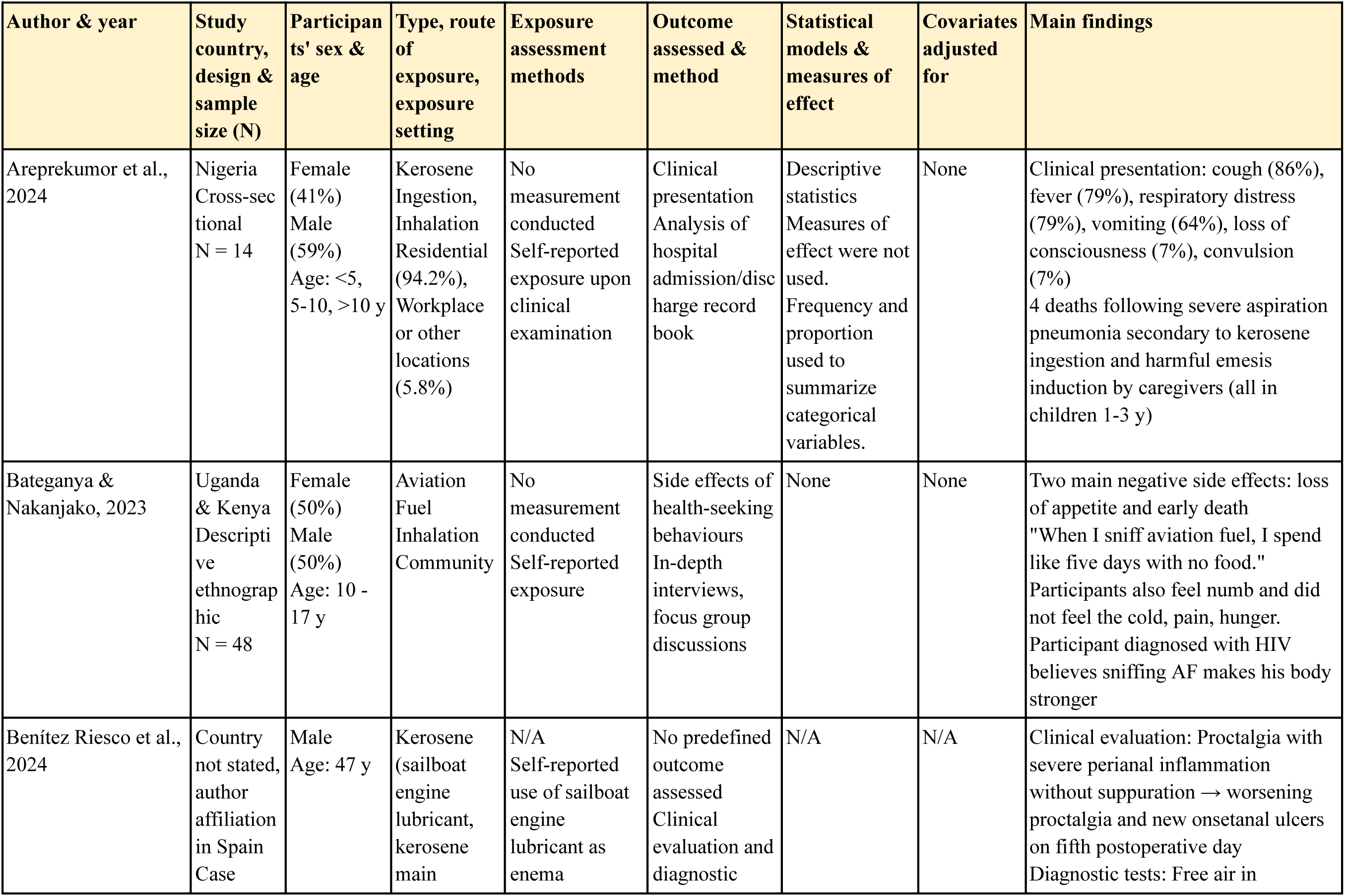
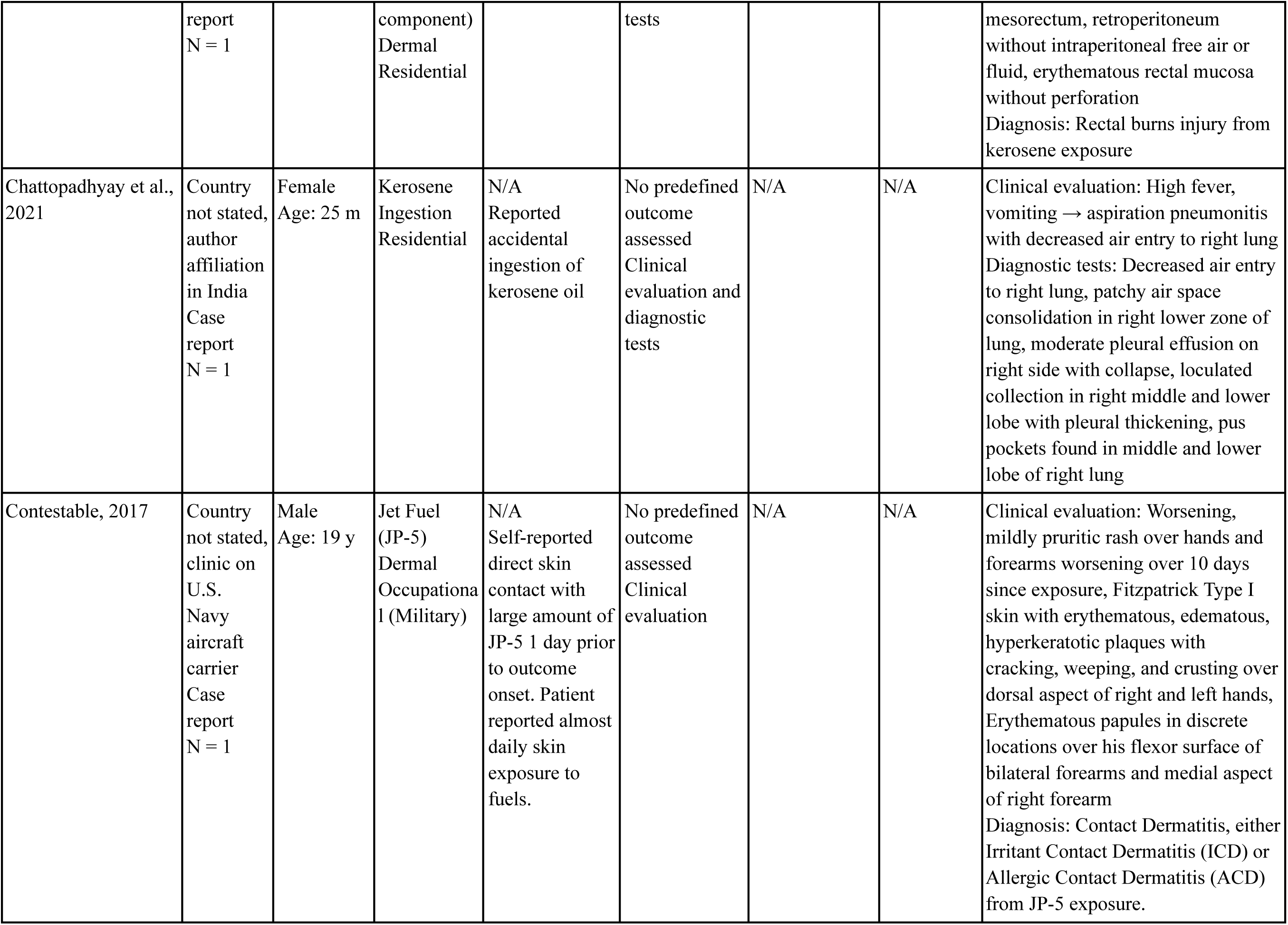
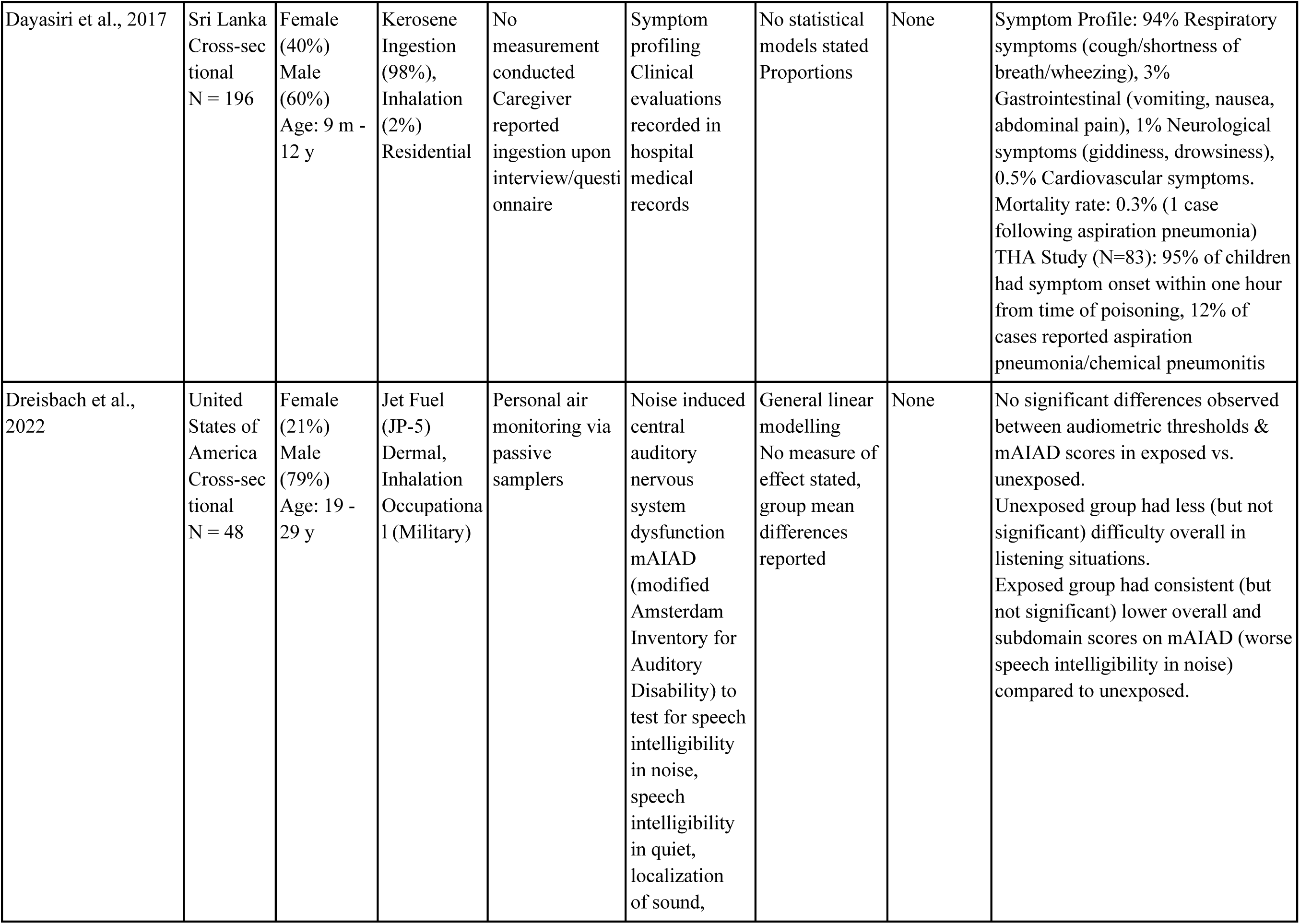
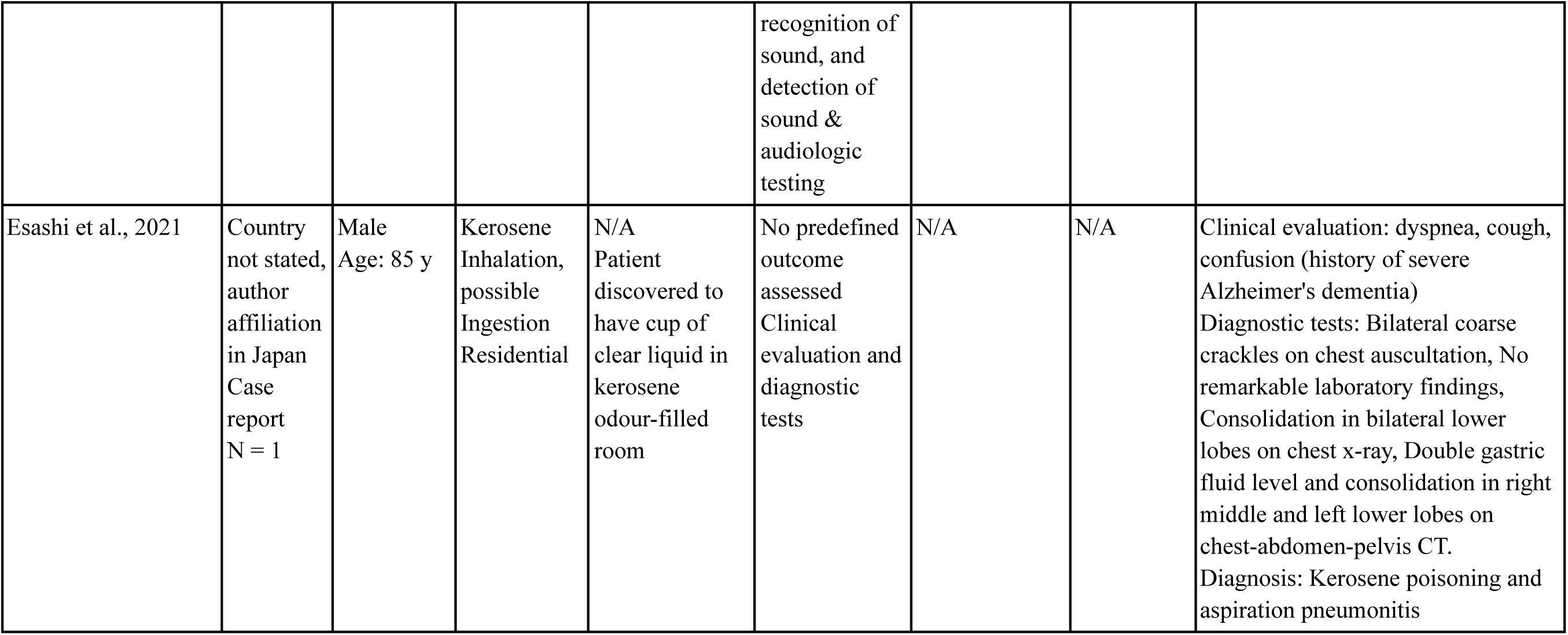
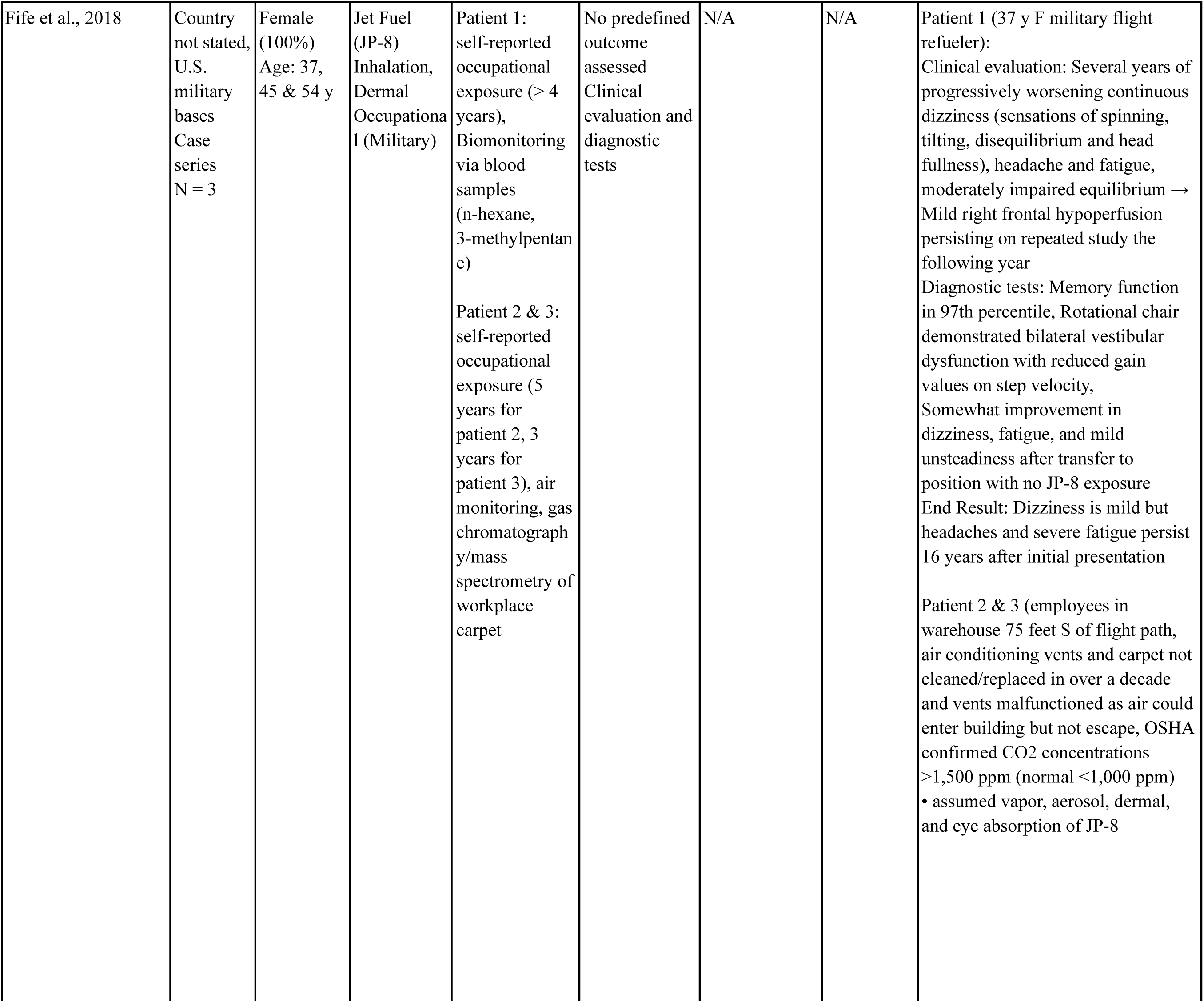
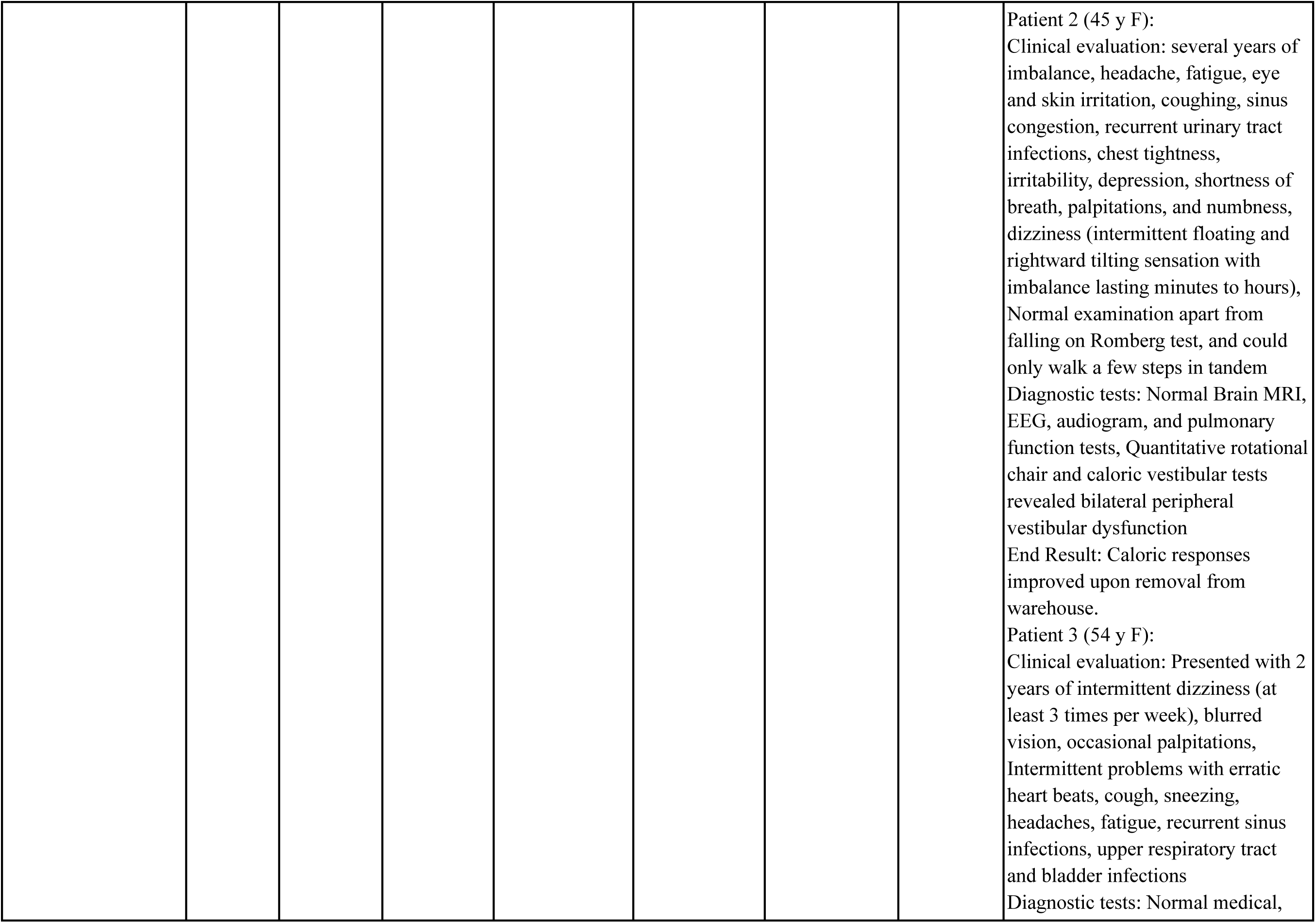
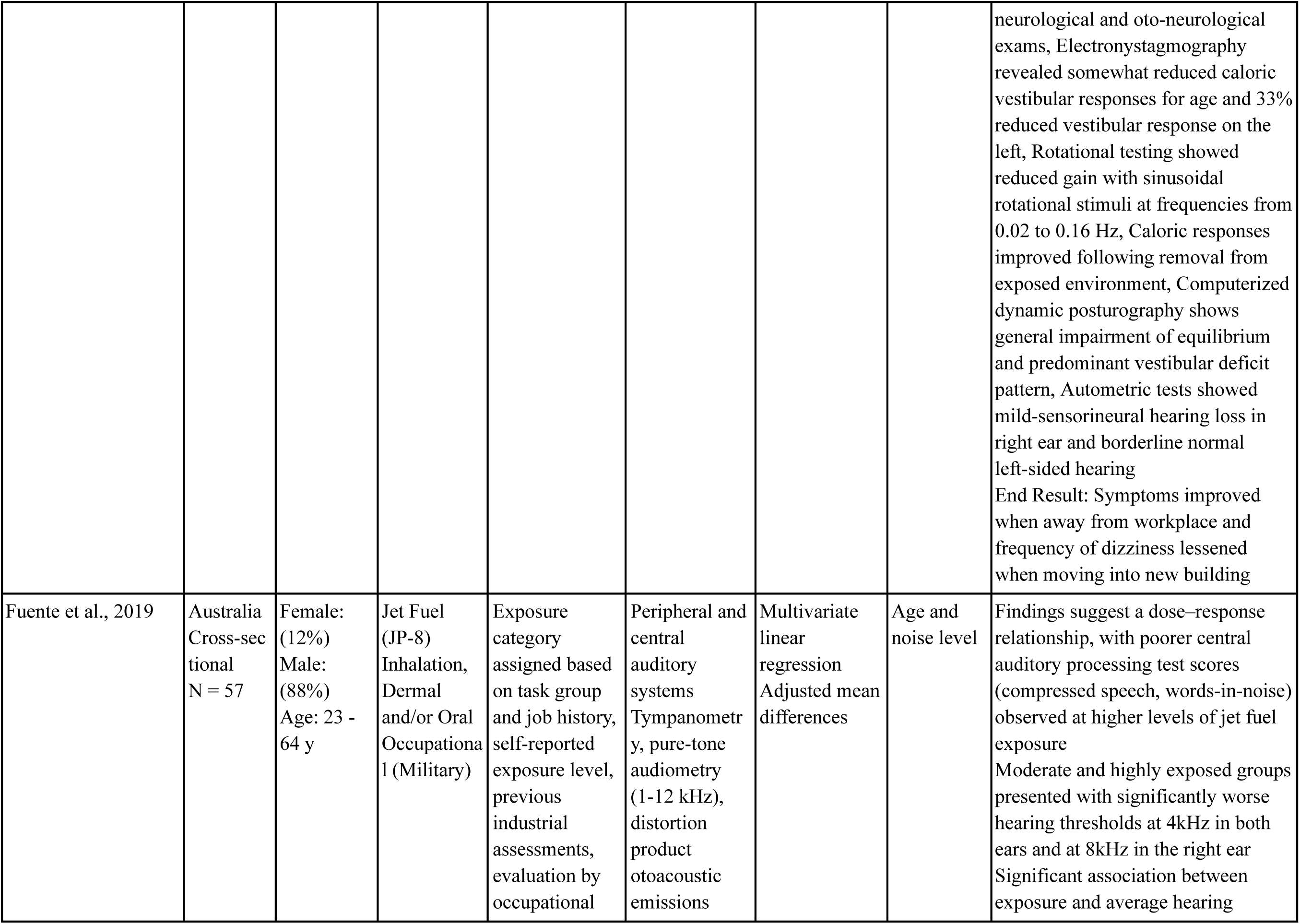
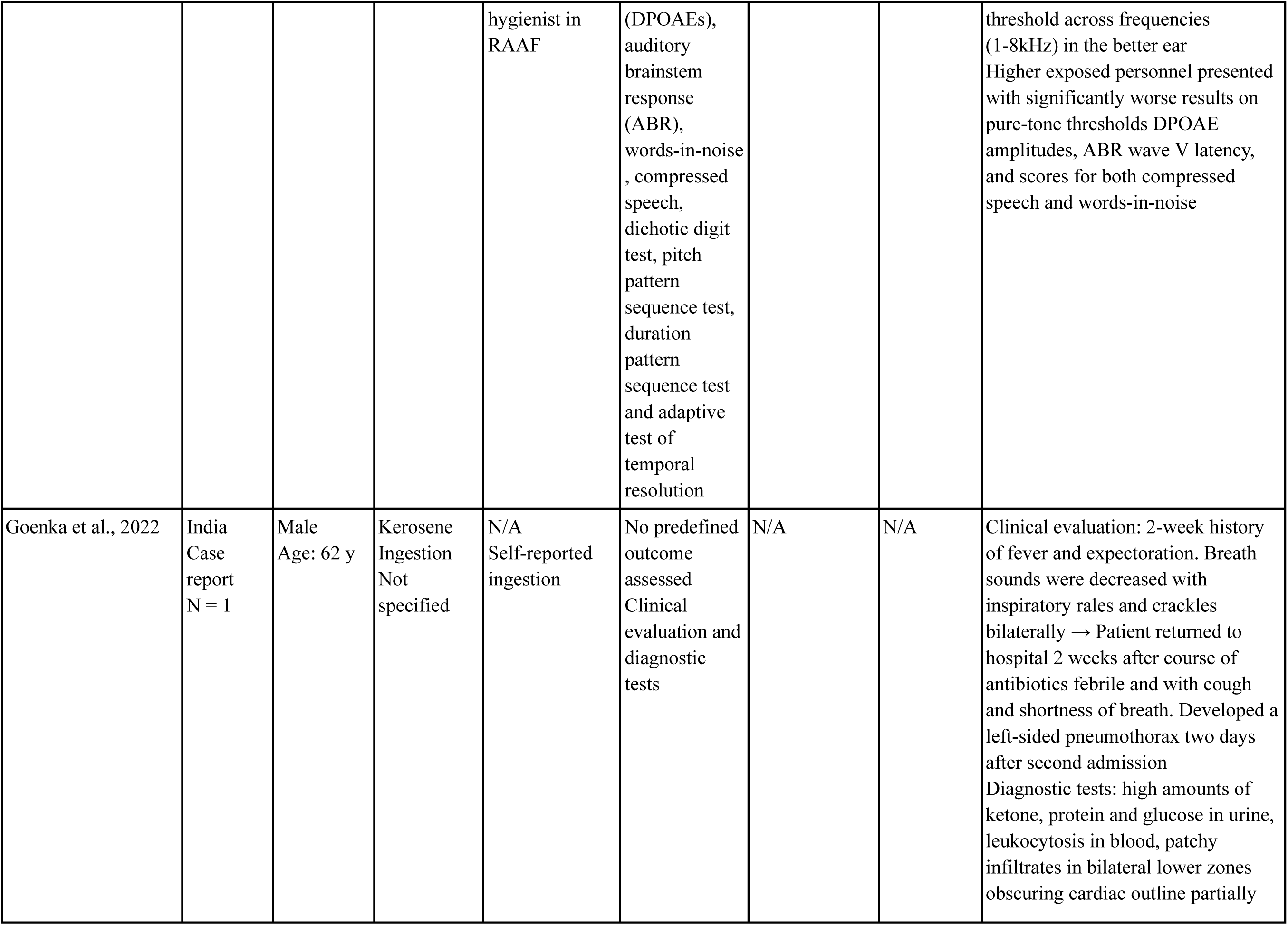
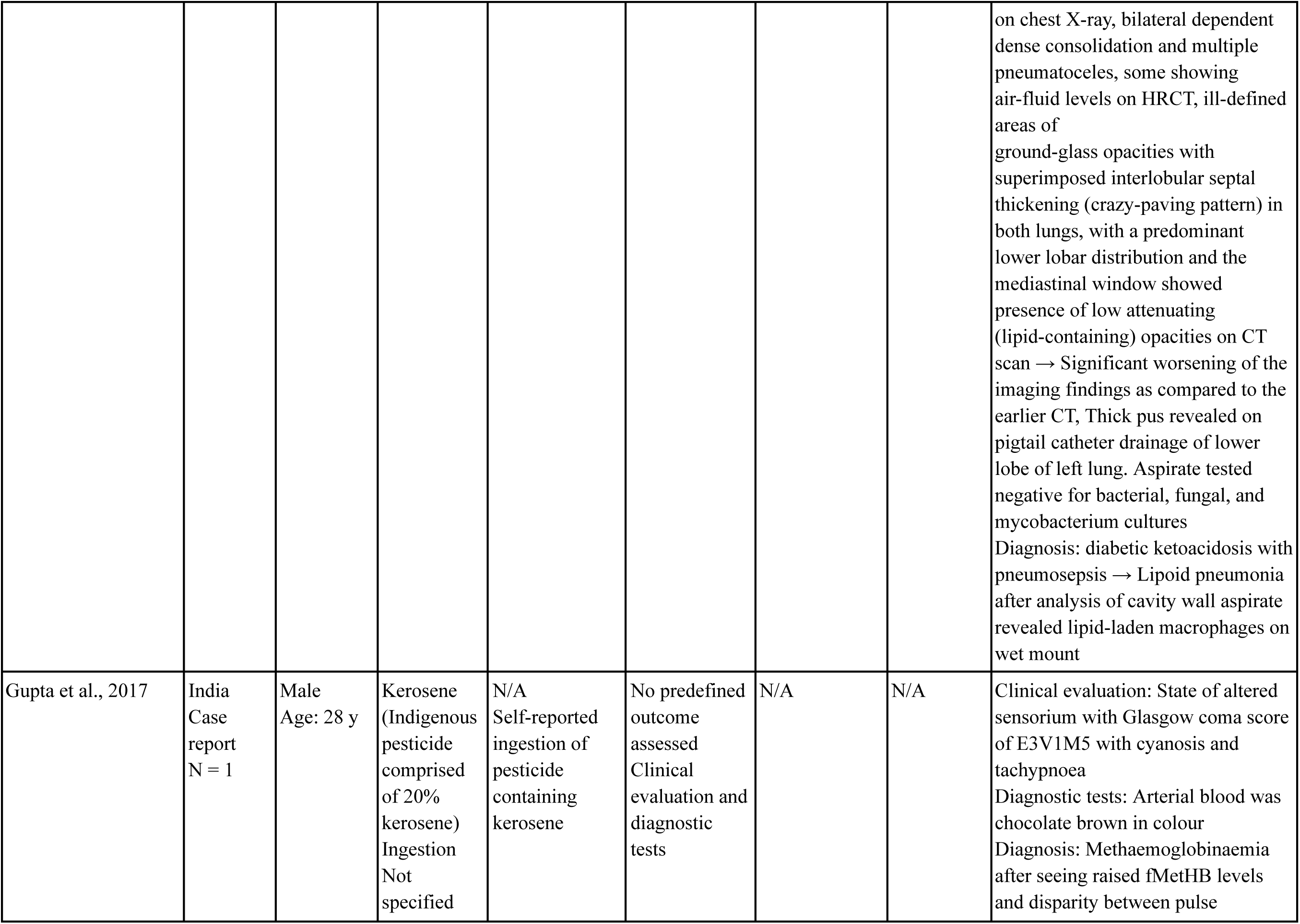
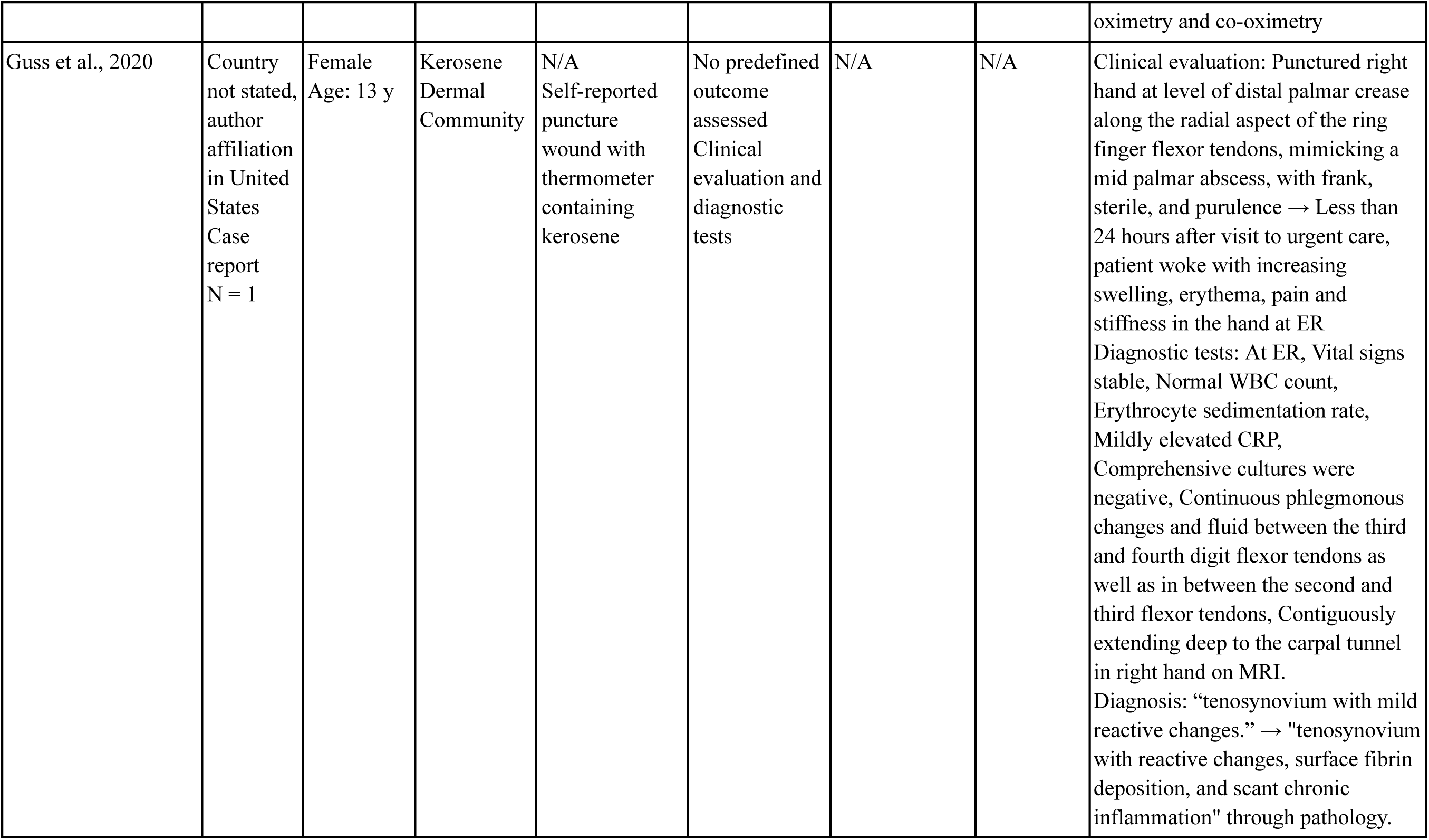
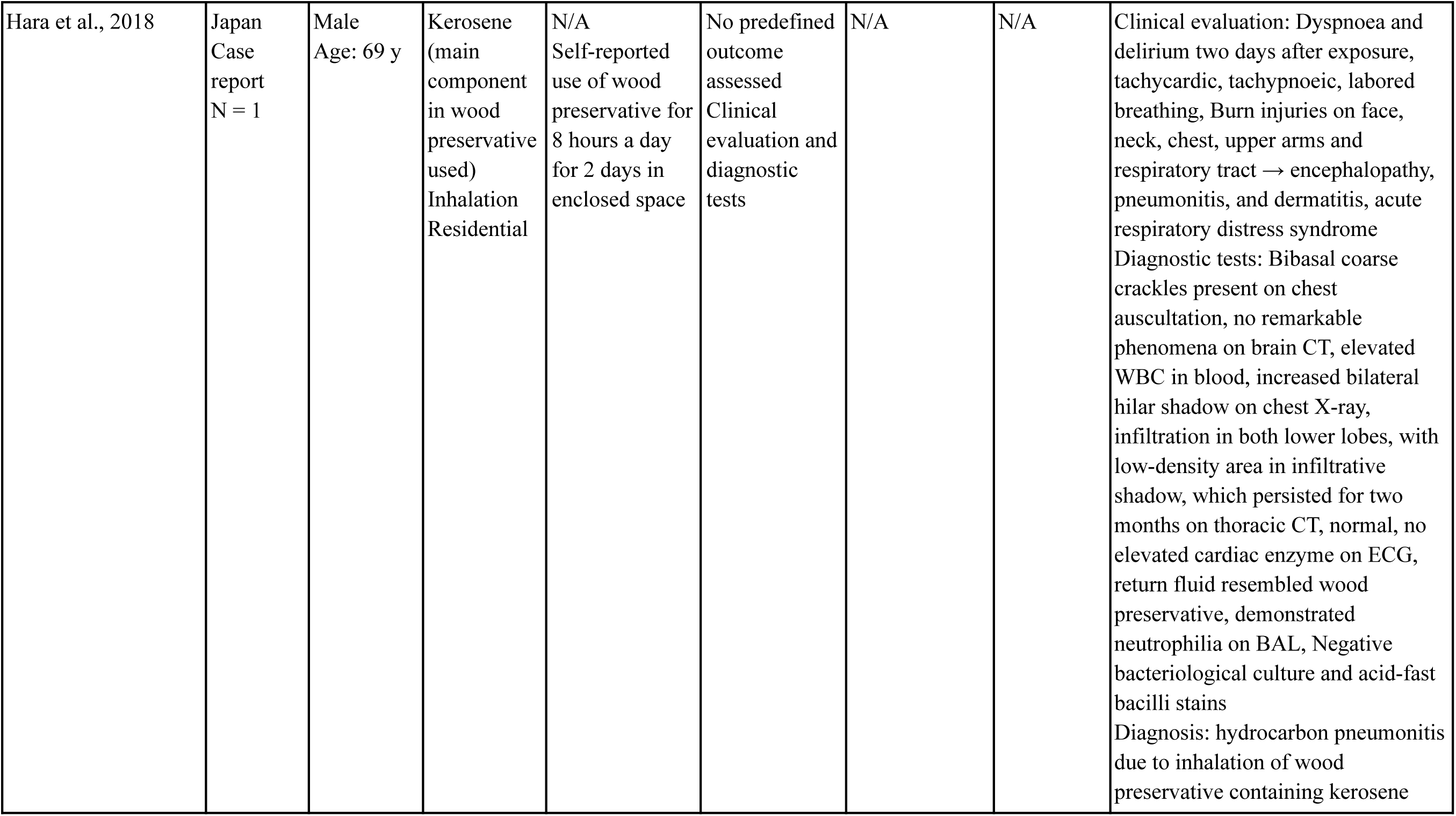
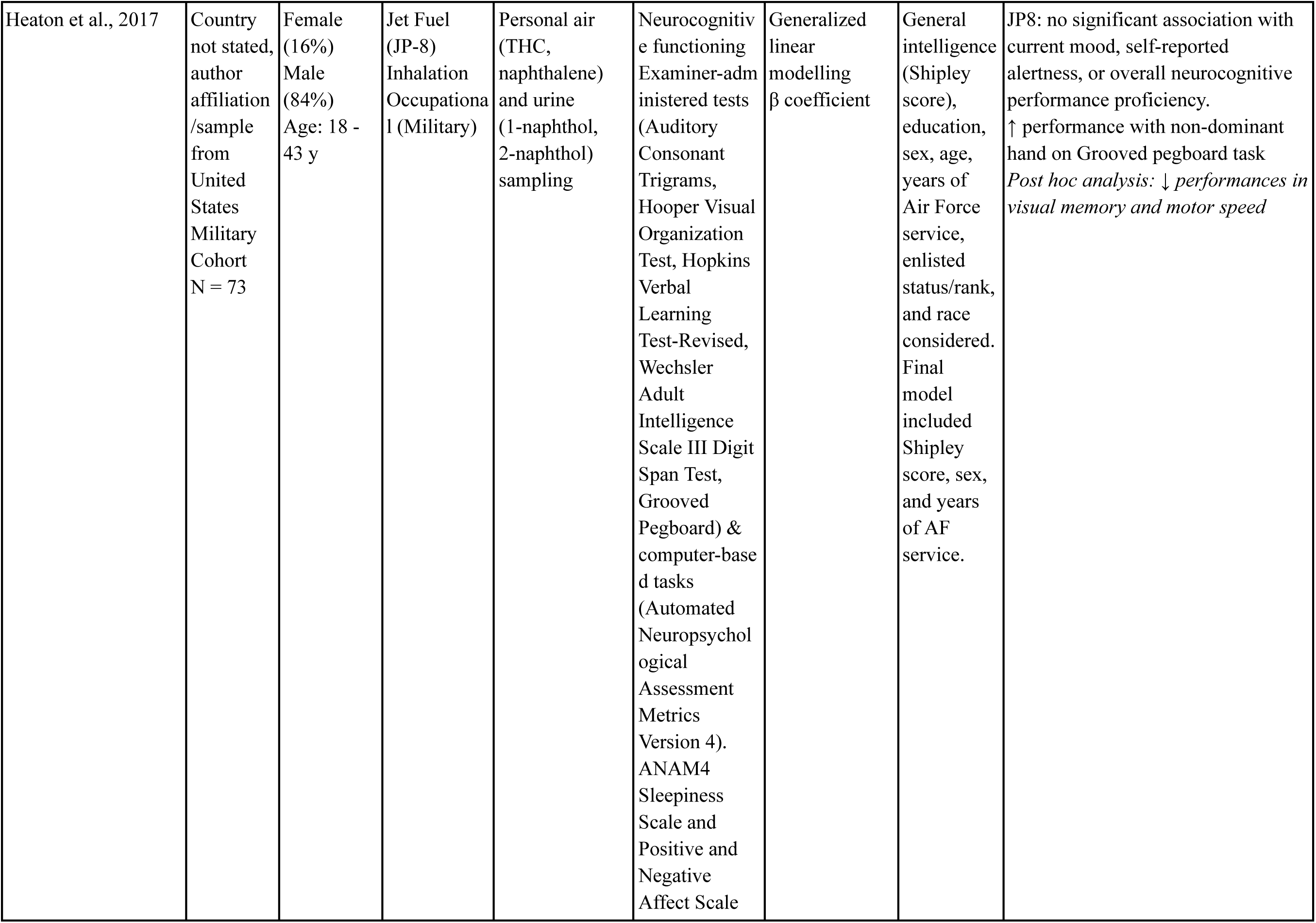
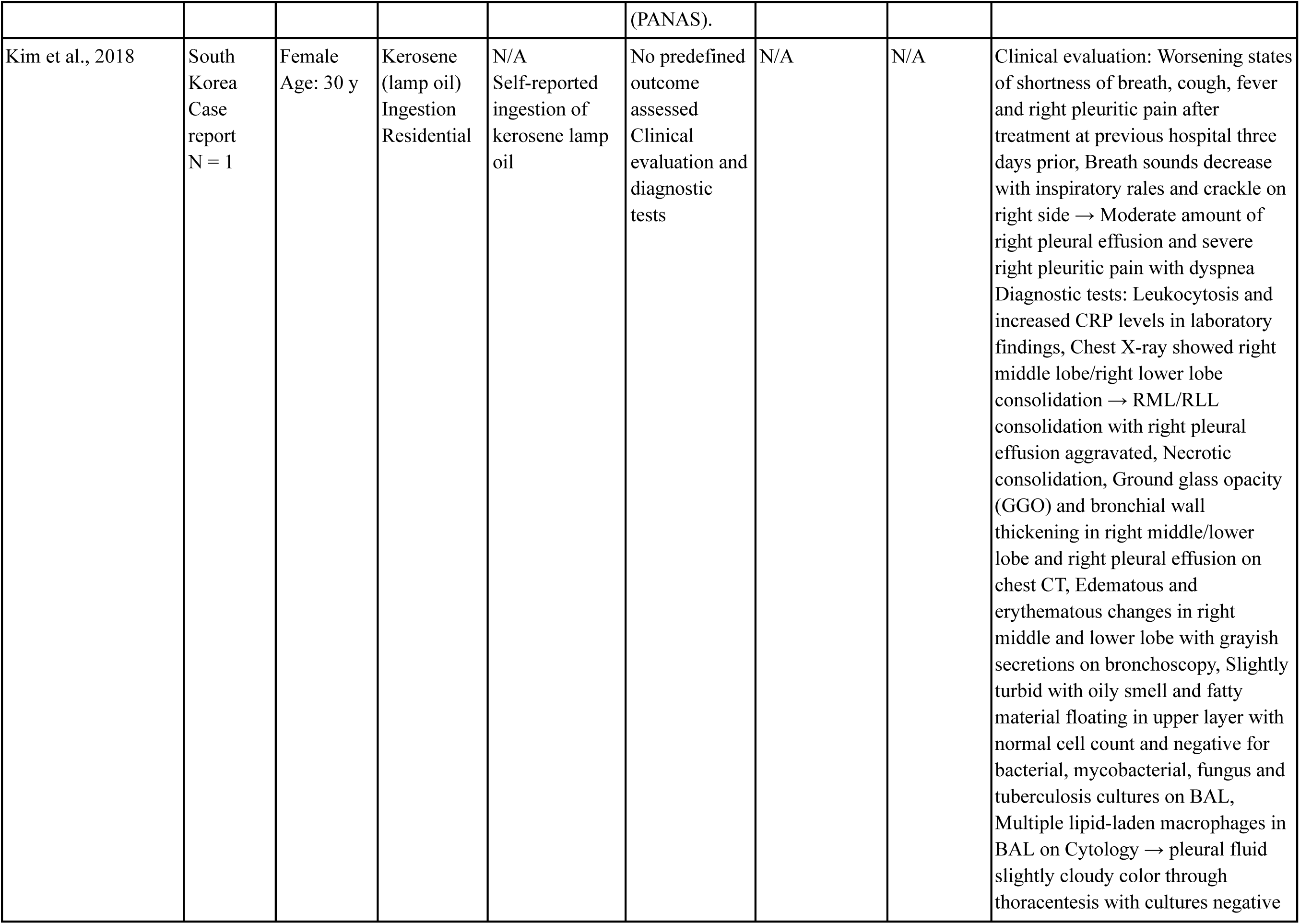
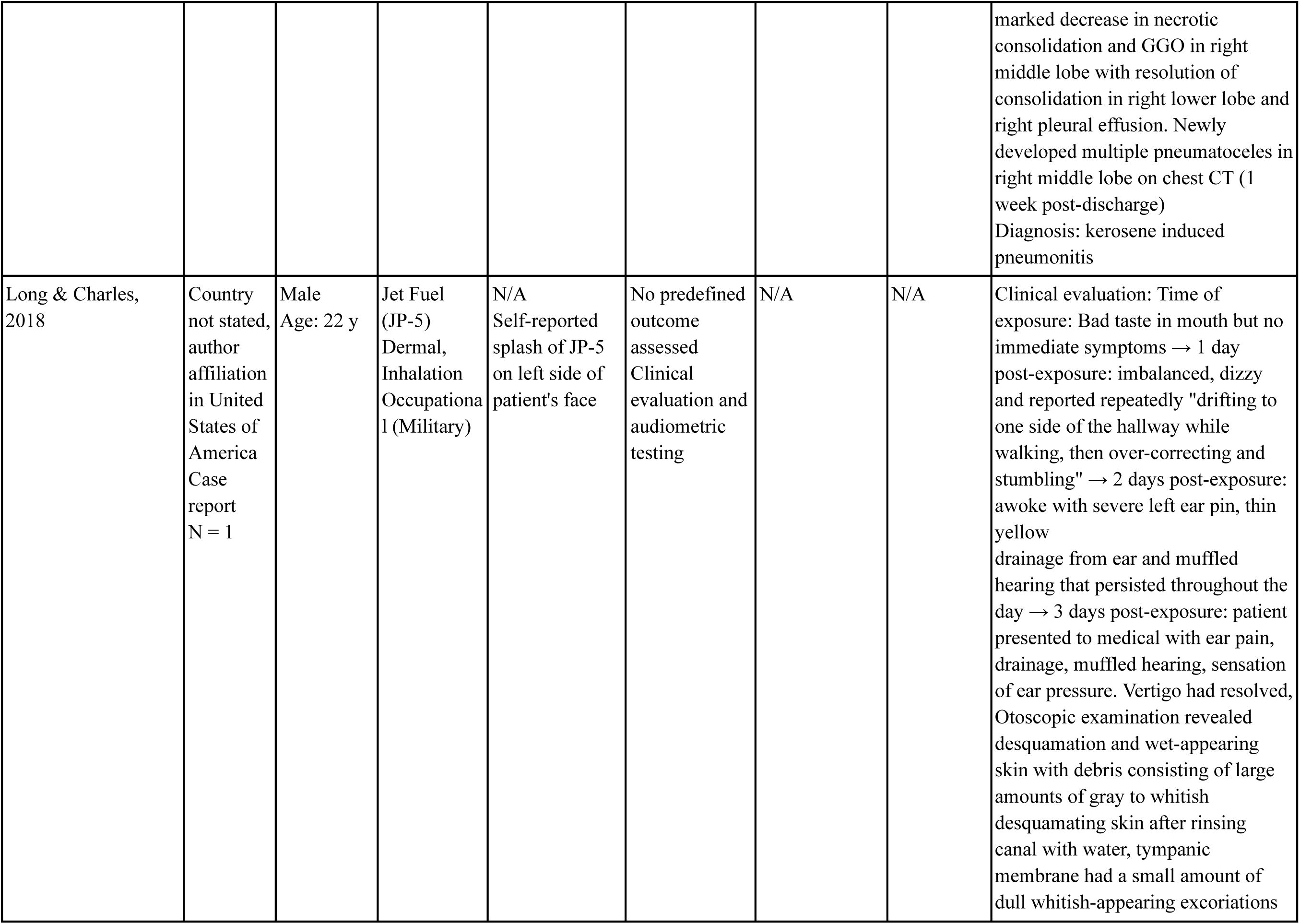
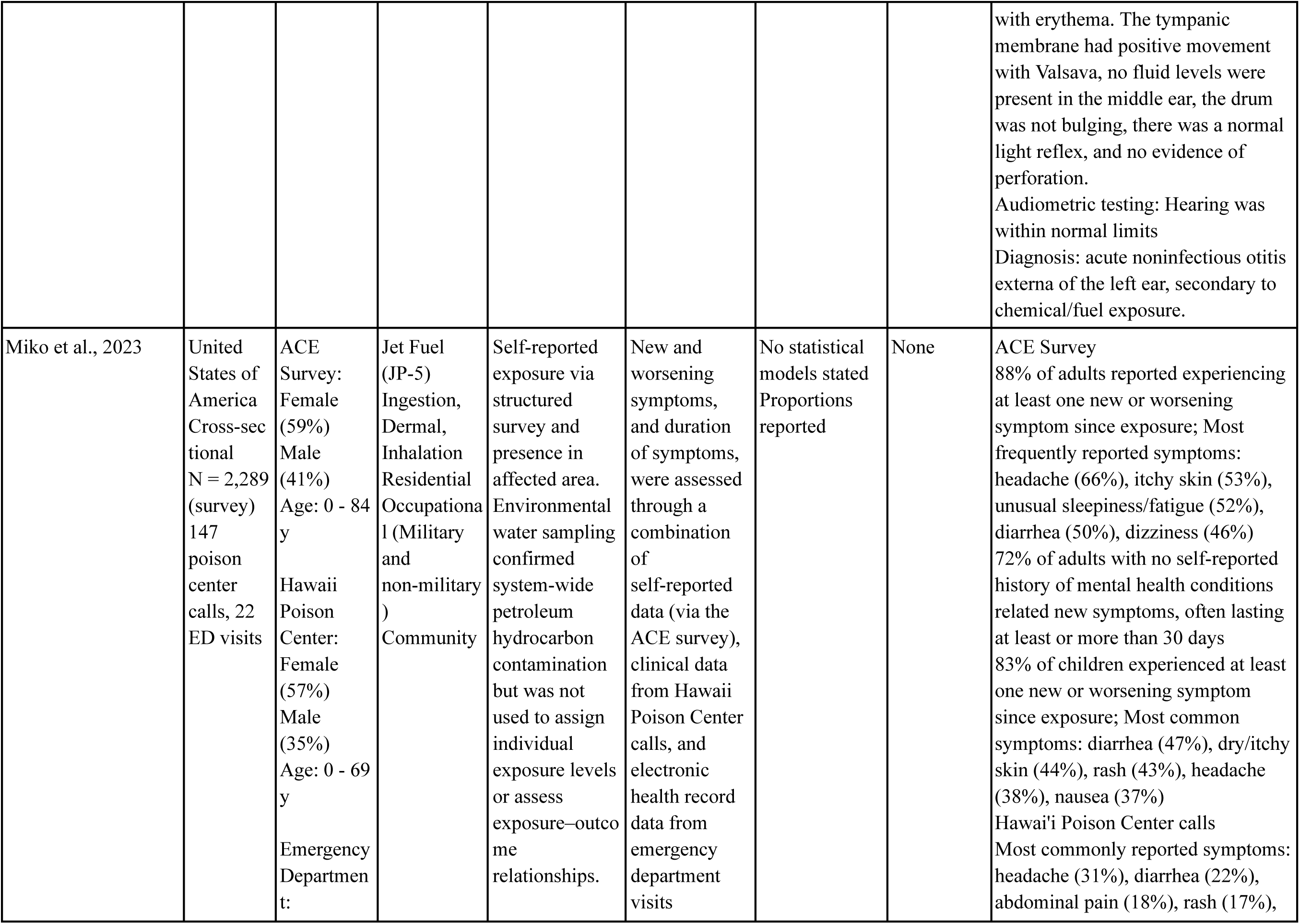
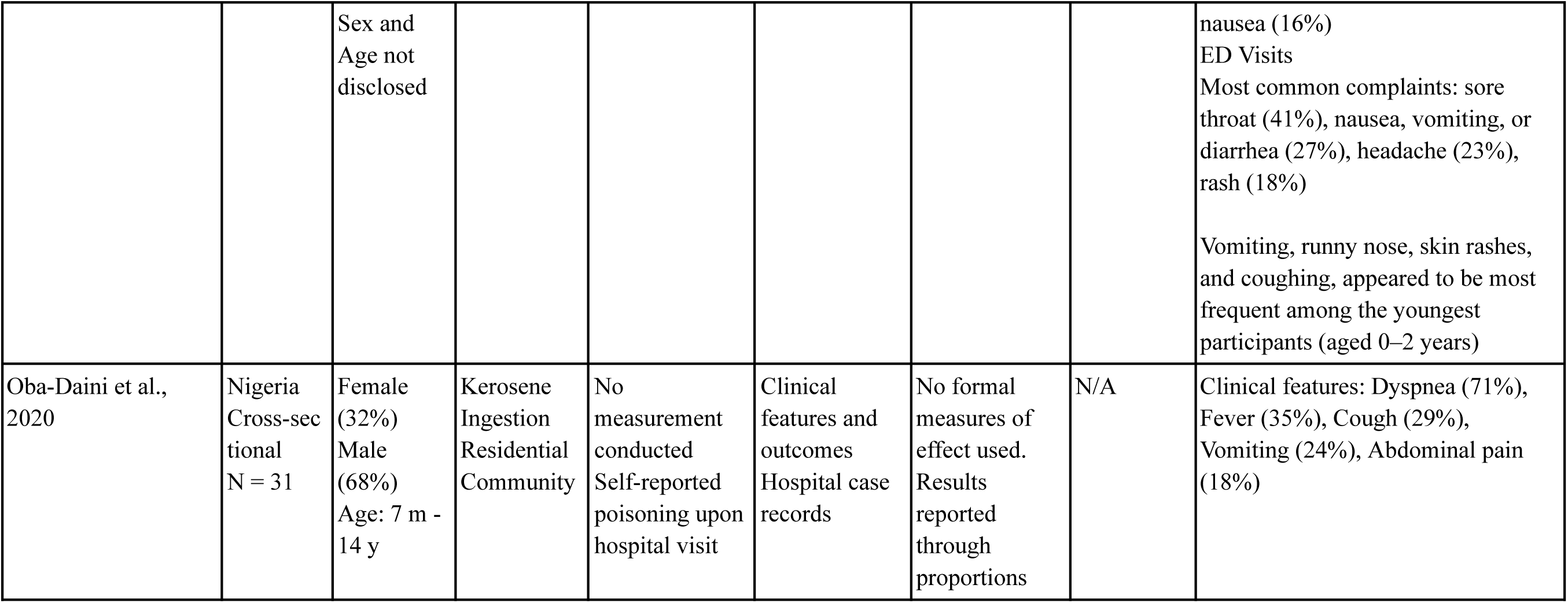
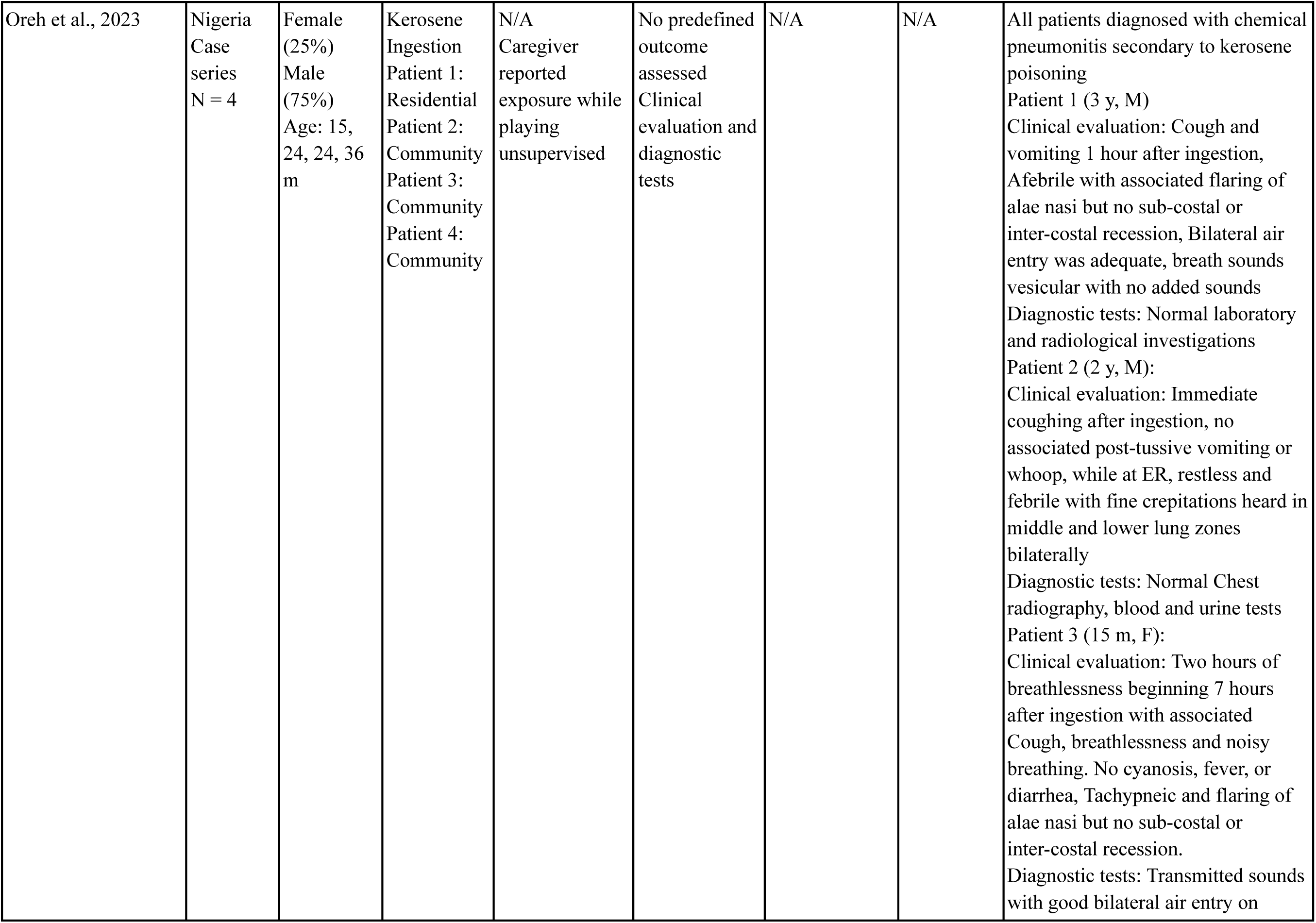
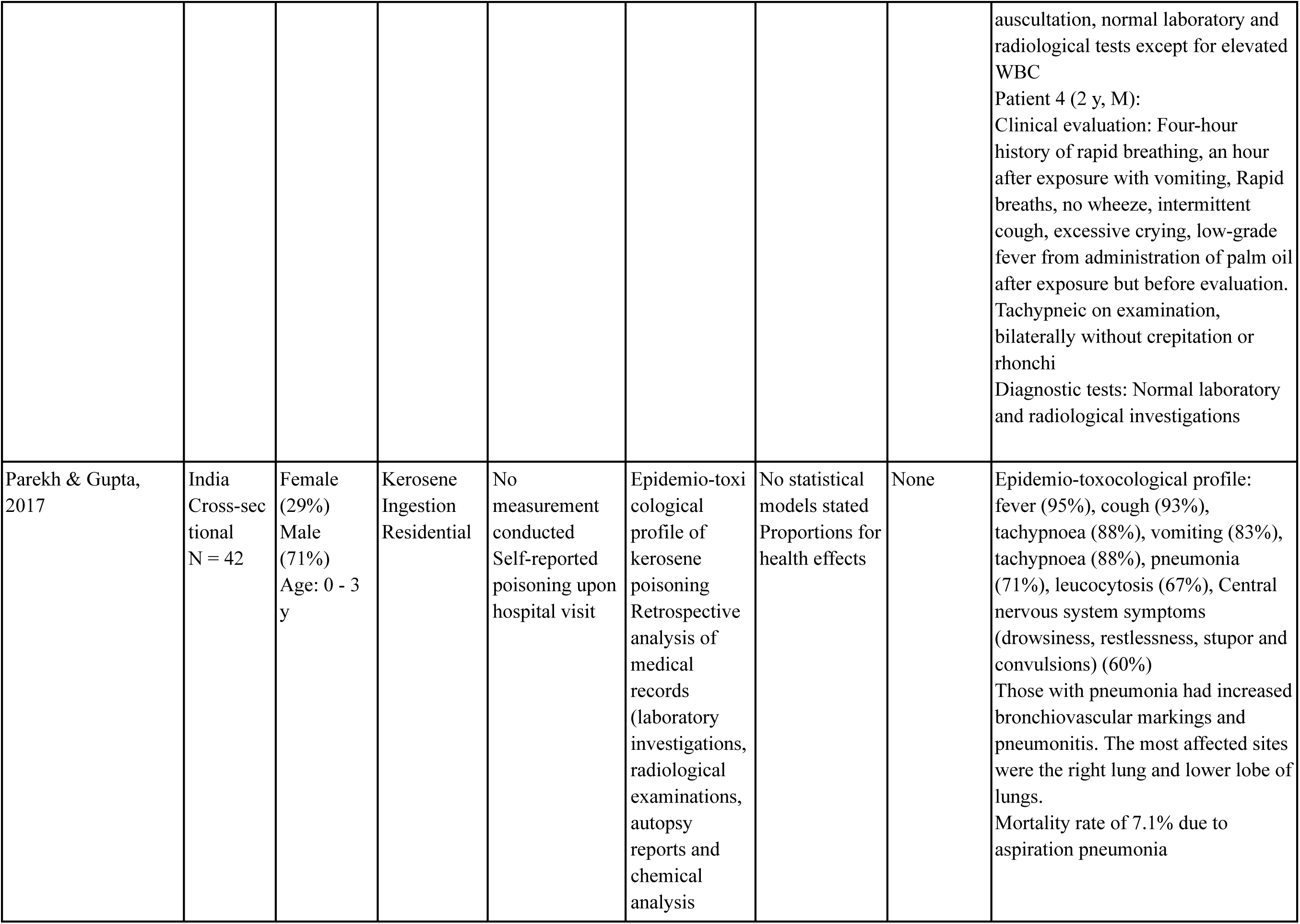
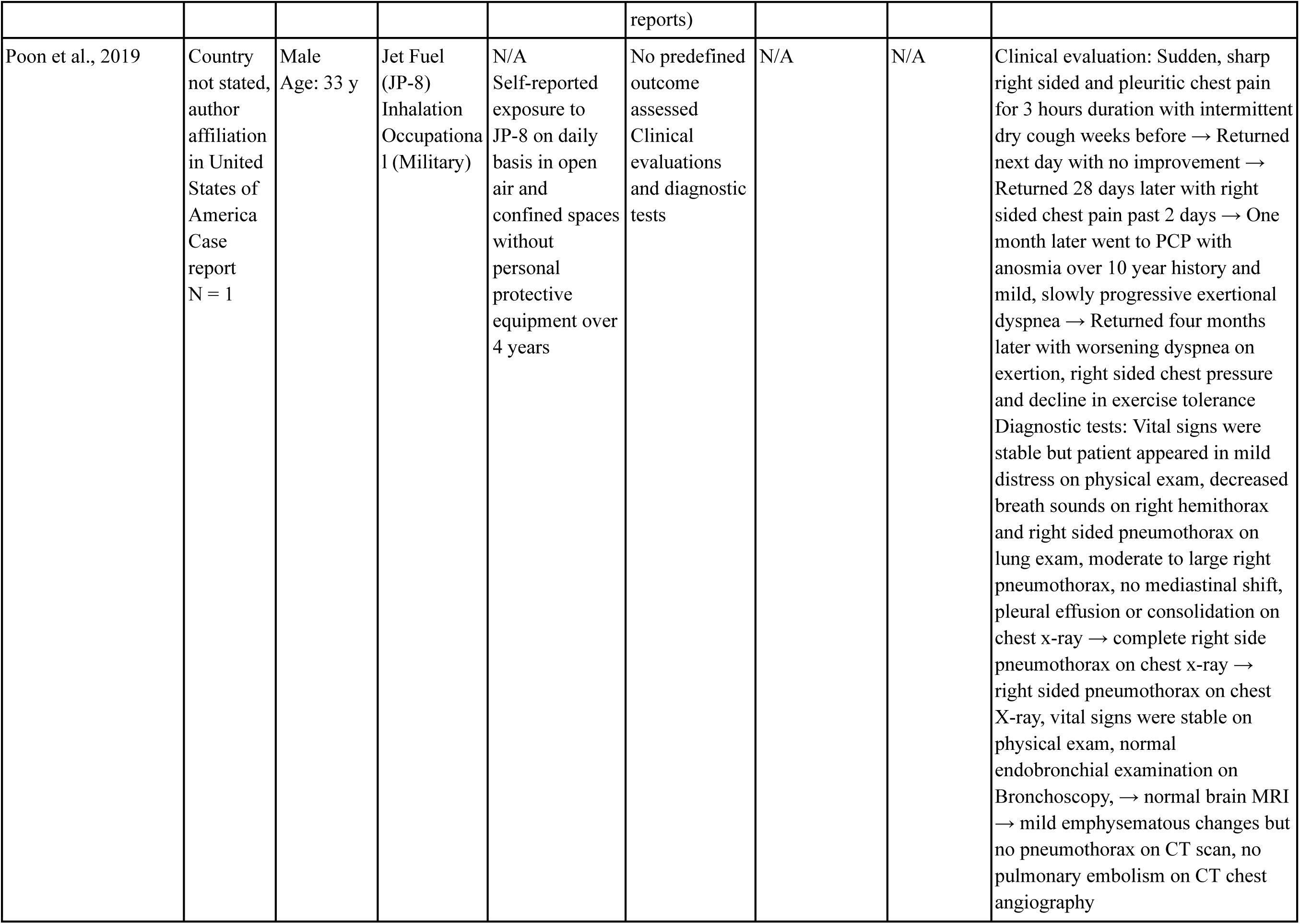
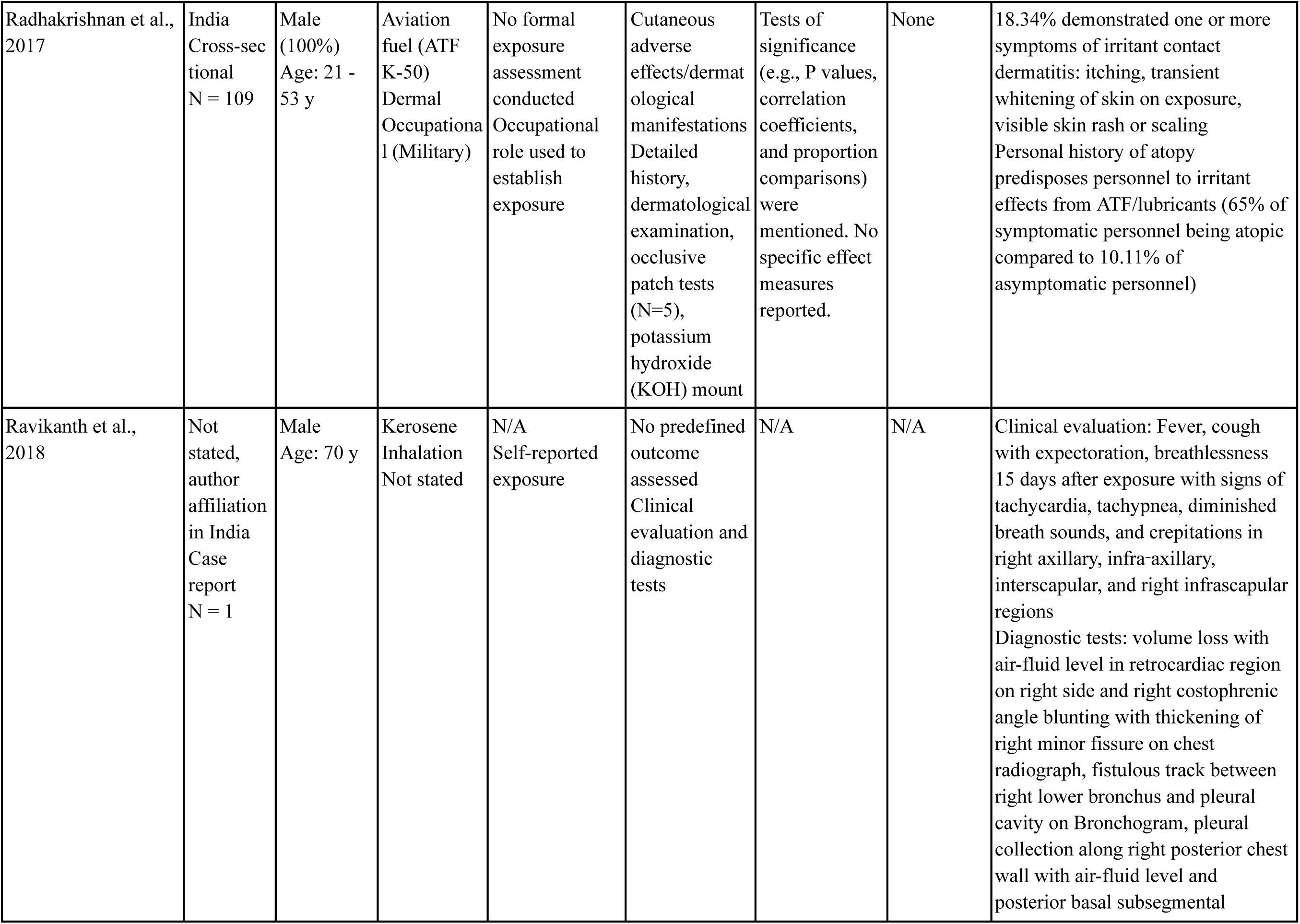
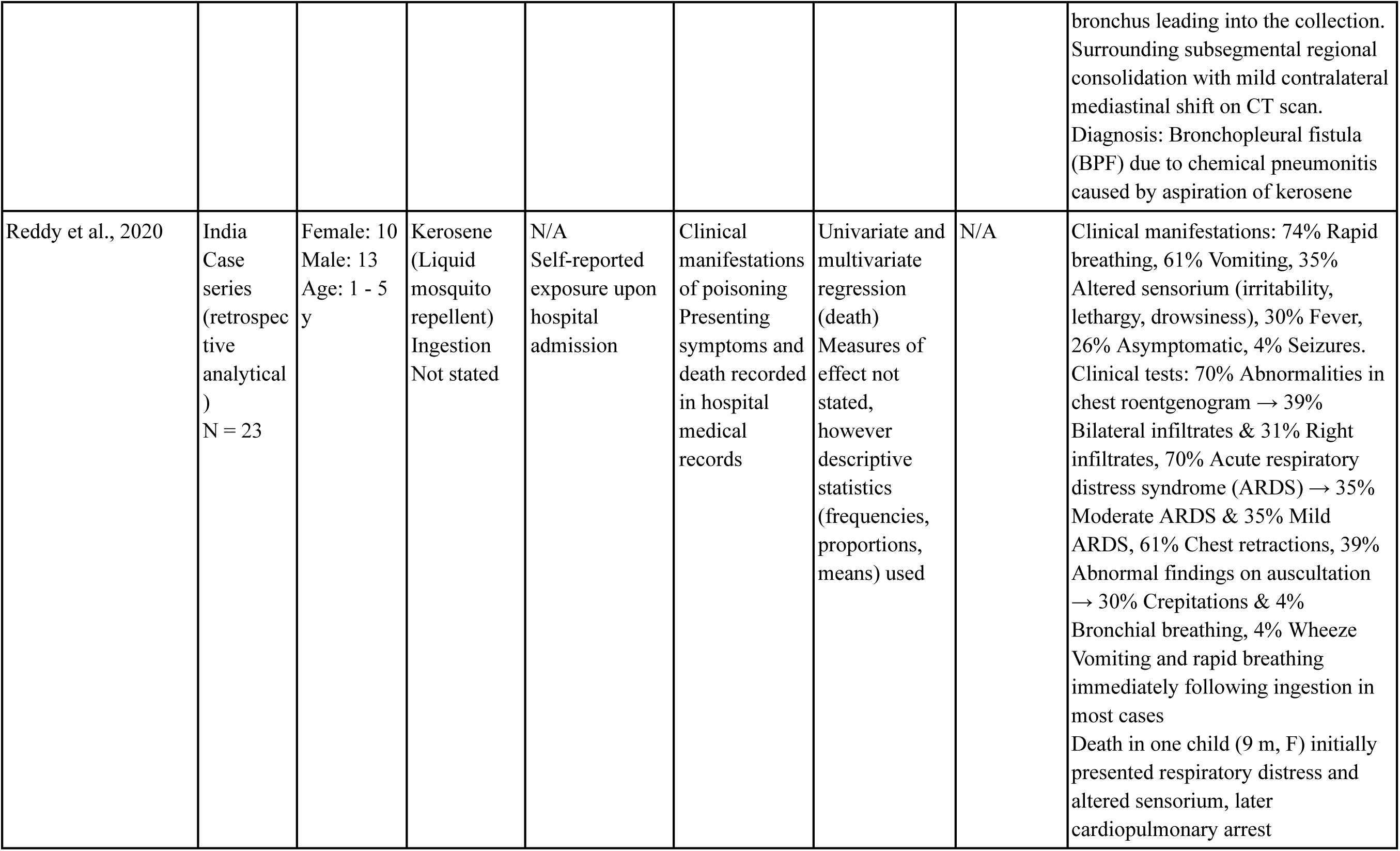
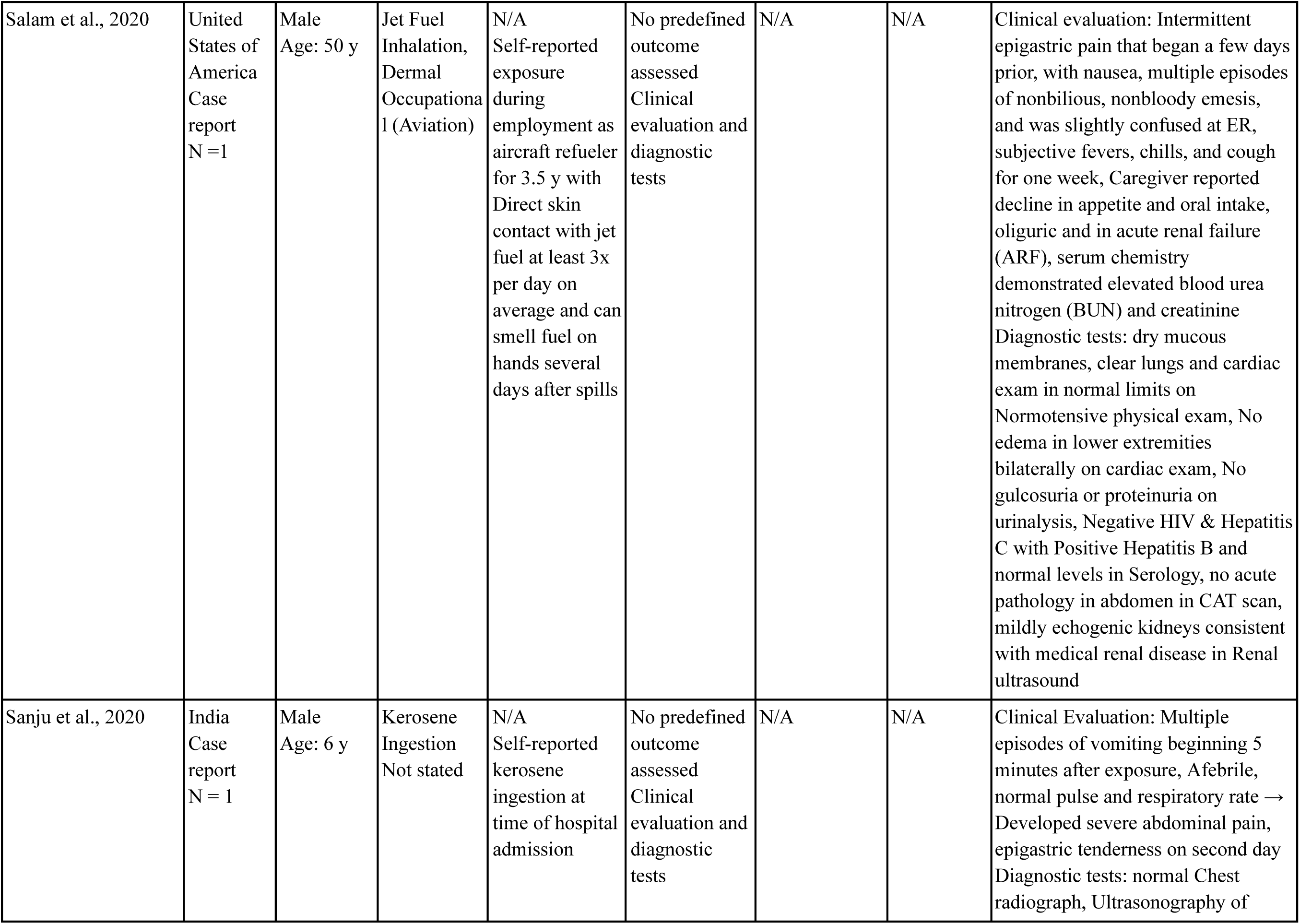
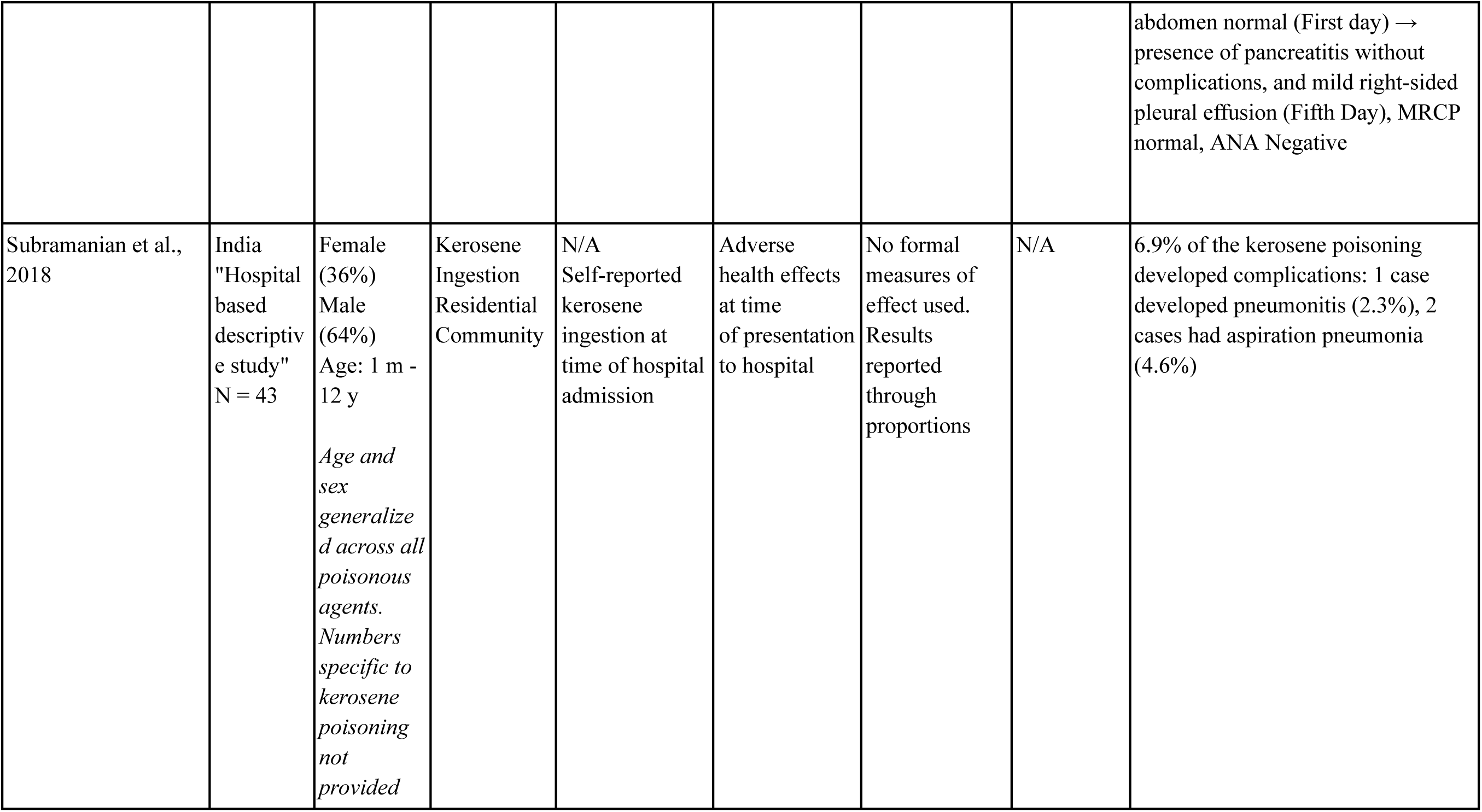
Data Extracted from Included Studies.

### 2.3 Quality Assessment

Four quality assessment checklists were developed with distinct considerations for the following study designs: trials, case-control/cohort studies, cross-sectional studies and case reports/case series, and can be found in Table S2. We adapted quality assessment criteria from the Modified Downs and Black Checklist, Joanna Briggs Checklist for Case Reports, and another systematic review in the field of environmental epidemiology (Carvajal et al., 2025; Downs & Black, 1998; Moola et al., 2020). To maintain consistency and transparency, each study was evaluated using the appraisal framework corresponding to its self-identified design. One study did not self-identify its design (Oba-Daini et al., 2020); however, given that their methods largely mirrored other descriptive cross-sectional studies identified in this review, the article was quality assessed against that framework. Quality assessment was conducted blindly and independently by two reviewers (VC and BN) for each included study. Total scores consisted of the combined scores of all indicators, which was then expressed as a percentage of the maximum score. Categories of quality were assigned based on the score in percentage: excellent quality (score ≥81%), good quality (between 61 and 80%), fair quality (between 41 and 60%), poor quality (between 21 and 40%) and very poor quality (≤20%).

While the criteria used in quality assessment do not necessarily reflect the overall quality of a given study, the information assessed is critical in the evaluation of the quality of evidence specific to the interest of this review. For instance, a paper may have incidentally reported findings pertaining to health effects of jet fuel and/or kerosene exposure, while not assessing this association as the primary interest of the study. These publications may be of high scientific quality independent of this review; however, the strength of evidence within the context of our research question may be limited. The results of these appraisals informed the synthesis and interpretation of the relationship between jet fuel and/or kerosene exposures on health outcomes reported within the included studies.

### 2.4 Analysis & Evaluation of Evidence

Observable and/or diagnosable health effects were prioritized, whereas laboratory or clinical measurements (e.g. respiratory rate, heart rate, oxygen saturation (SpO_2_)) were excluded from analyses. To facilitate synthesis and analysis, all reported symptoms were extracted and categorized. Within each bodily system, raw symptoms and diagnoses were reviewed and organized into symptom categories (e.g. “dyspnea”, “shortness of breath” and “rapid breathing” were grouped under category “Laboured breathing” within the Respiratory system). Binary coding was conducted for each study, with a value of 1 assigned when authors reported at least one symptom in the corresponding category. Frequency counts were then used to assess the distribution of symptoms across studies, and stratified by exposure type (kerosene or jet fuel), and route (oral, dermal, or inhalation) when possible. Full mappings of extracted health effects and categories can be found in Table S3.

The quality assessment scores account for any design and methodological limitations of a given study, and were used to contextualize the strength of evidence in our synthesis and evaluation of reported findings. A subset analysis limited to good and excellent quality studies was conducted where frequency counts were reassessed to evaluate health effects supported by higher quality evidence. These patterns were then compared to those observed across all included studies to evaluate consistency of findings.

## 3. Results

### 3.1. Selected Studies

A total of 2,216 articles were retrieved from the search. After deduplication, title and abstract screening, 72 articles remained for full text review. One additional eligible article was found upon retrospective citation tracing. Ultimately, 28 articles met all eligibility criteria and were included in this review. The PRISMA flow diagram outlining the article selection process is presented in Figure 1.

### 3.2 Characteristics of Included Studies

#### 3.2.1. Study Design

All included studies were observational in design; no randomized controlled trials or other intervention studies were identified. The vast majority of included studies (89%) were descriptive in nature and did not conduct formal exposure assessment or compare outcomes by exposure level, including case reports (N=14), cross-sectional studies (N=6), case series (N=3), one “hospitable based descriptive study” and one ethnographic study. Three analytical studies (11%) were identified in this review: two cross-sectional studies (Dreishbach et al., 2022; Fuente et al., 2019) and one cohort study (Heaton et al., 2017). Each of these utilized comparison groups to test relationships between exposure to JP-5 or JP-8 and human health outcomes.

#### 3.2.2. Participant Demographics

Males were represented in 23 (85%) studies, compared to females who were assessed in 15 (56%) and comprised at least 50% of the sample in only 6 (22%) studies. The age group most observed across included articles were adults of working age (aged 18 to 64 years) (N=14), followed by children (aged 2 to 17 years) (N=12), infants (up to and including one year) (N=7) and older adults (aged 65 and older) (N=4). However, in kerosene-specific studies, children (N=10, 59%) and infants (N=6, 35%) were most assessed, compared to those specific to jet fuel in which the vast majority of reports assessed adults of working age (N=10, 91%). Adults aged 65 years and older were the least assessed group across all included studies (N=4). Sample sizes ranged from single-patient case reports to a survey of 2,289 participants (Miko et al., 2023). A summary of participant age groups by fuel type is presented in Table 3.

**Table 3.** Characteristics of Included Studies.

| **Characteristic** | **Kerosene (N=17) (%)** | **Jet Fuel (N=11) (%)** |
| --- | --- | --- |
| **Exposure Route** |  |  |
| Oral | 12 (71%) | 2 (18%) |
| Inhalation | 5 (29%) | 9 (82%) |
| Dermal | 2 (12%) | 8 (73%) |
| **Exposure Duration** |  |  |
| Acute | 17 (100%) | 1 (9%) |
| Chronic | 0 | 10 (91%) |
| **Exposure Setting** |  |  |
| Residential | 11 (65%) | 1 (9%) |
| Community | 4 (24%) | 2 (18%) |
| Occupational (Non-military & military) | 1 (6%) | 10 (91%) |
| Occupational (Non-military) | 1 (6%) | 2 (18%) |
| Occupational (Military) | 0 | 9 (82%) |
| Not stated | 5 (29%) | 0 |
| **Age Group** |  |  |
| Children | 10 (59%) | 2 (18%) |
| Infants | 6 (35%) | 1 (9%) |
| Adults | 4 (24%) | 10 (91%) |
| Older Adults | 3 (18%) | 1 (9%) |

*\*Number (N) and proportion (%) of studies reporting characteristic*

#### 3.2.3. Country and Year of Publications

Kerosene-specific reports appeared consistently throughout the review period (2017-2024), with the exclusion of the year 2019, and were published primarily in low- and middle-income countries, including India (N=8) and Sri Lanka (N=1). Other kerosene-specific reports originated from Japan (N=2), South Korea, Spain and the United States of America (N=1 each). In contrast, jet fuel-specific articles originated from higher-income countries primarily, including the United States of America (N=8) and Australia (N=1). Few studies were also published in Kenya and Uganda (N=1) and India (N=1). Only 3 (27%) jet fuel-specific studies were published after 2020, with none identified in the years 2021 or 2024.

### 3.3. Quality Assessment

Average quality scores varied by study design, with case reports scoring highest on average at 77% (“good” quality). These publications typically scored well on items regarding descriptions of patients and their presenting conditions, but scored poorly when assessed for reporting of unanticipated or harmful events following clinical treatment. Cross-sectional studies were generally of “fair” quality with an average score of 49%. Common limitations of these studies included insufficient, or lack of: reporting on participant recruitment and demographic characteristics, consideration of confounders, formal exposure assessment, as well as analyses needed to draw dose-response conclusions about the associations of interest for this review. The sole cohort study identified in this review ranked “good” quality with a score of 71%. Notably, two of the three analytical studies identified by this review scored of higher quality (good or excellent) (Fuente et al., 2019; Heaton et al., 2017). Only one study scored of poorer quality. A summary of quality assessment results can be found in Table 4. Due to the absence of comparable studies, quality assessment was not conducted on one descriptive ethnographic study (Bateganya & Nakanjako, 2023), one “hospital based descriptive study” (Subramanian et al., 2018) and one analytical case series (Reddy et al., 2020). A full breakdown of quality assessment items for each study can is presented in Table S4-S6.

**Table 4.** Results of Quality Assessment.

| Author & Year | Study Design | Score (%) | Score Quality Category |
| --- | --- | --- | --- |
| Kim et al., 2018 | Case report | 90 | Excellent |
| Long & Charles, 2018 | Case report | 90 | Excellent |
| Poon et al., 2019 | Case report | 90 | Excellent |
| Benítez Riesco et al., 2024 | Case report | 70 | Good |
| Chattopadhyay et al., 2021 | Case report | 80 | Good |
| Contestable, 2017 | Case report | 80 | Good |
| Esashi et al., 2021 | Case report | 70 | Good |
| Fuente et al., 2019 | Cross-sectional | 71 | Good |
| Goenka et al., 2022 | Case report | 80 | Good |
| Gupta et al., 2017 | Case report | 70 | Good |
| Guss et al., 2020 | Case report | 70 | Good |
| Hara et al., 2018 | Case report | 80 | Good |
| Heaton et al., 2017 | Cohort | 70 | Good |
| Oreh et al., 2023 | Case report | 80 | Good |
| Ravikanth et al., 2018 | Case report | 80 | Good |
| Salam et al., 2020 | Case report | 70 | Good |
| Sanju et al., 2020 | Case report | 80 | Good |
| Areprekumor et al., 2024 | Cross-sectional | 43 | Fair |
| Dayasiri et al., 2017 | Cross-sectional | 48 | Fair |
| Dreisbach et al., 2022 | Cross-sectional | 52 | Fair |
| Fife et al., 2018 | Case report | 50 | Fair |
| Miko et al., 2023 | Cross-sectional | 48 | Fair |
| Parekh & Gupta, 2017 | Cross-sectional | 48 | Fair |
| Radhakrishnan et al., 2017 | Cross-sectional | 48 | Fair |
| Oba-Daini et al., 2020 | Cross-sectional | 38 | Poor |
| Bateganya & Nakanjako, 2023* | Ethnographic study | N/A | N/A |
| Reddy et al., 2020* | Analytical case series | N/A | N/A |
| Subramanian et al., 2018* | Hospital-based descriptive study | N/A | N/A |

\*Quality assessment not conducted due to lack of comparison studies

### 3.4. Characteristics of Exposure

A summary of exposure durations, settings and routes by fuel type is presented in Table 3.

#### 3.4.1. Exposures and Durations Assessed

Jet fuel-specific studies comprised 39% of included publications (N=11) (Bateganya & Nakanjako, 2023; Contestable, 2017; Dreisbach et al., 2022; Fife et al., 2018; Fuente et al., 2019; Heaton et al., 2017; Long & Charles, 2018; Miko et al., 2023; Poon et al., 2019; Radhakrishnan et al., 2017; Salam et al., 2020). Among these, JP-5 (N=4) and JP-8 (N=4) were most frequently assessed. Other less reported fuels included JP-4 or generalized jet/aviation fuel without specification of subtypes. Of these 11 jet fuel specific-studies, six (55%) scored as good or excellent quality (Contestable, 2017; Fuente et al., 2019; Heaton et al., 2017; Long & Charles, 2018; Poon et al., 2019; Salam et al., 2020). The greater part of all jet-fuel specific studies involved chronic occupational exposures (N=10, 91%).

In contrast, kerosene-specific reports comprised the larger portion of included studies (N=17, 61%), all of which involved acute exposure events (Areprekumor et al., 2024; Benítez Riesco et al., 2024; Chattopadhyay et al., 2021; Dayasiri et al., 2017; Esashi et al., 2021; Goenka et al., 2022; Gupta et al., 2017; Guss et al., 2020; Hara et al., 2018; Kim et al., 2018; Oba-Daini et al., 2020; Oreh et al., 2023; Parekh & Gupta, 2017; Ravikanth et al., 2018; Reddy et al., 2020; Sanju et al., 2020; Subramanian et al., 2018). Five of these studies reported on health effects of kerosene-containing products, including lamp oil, sailboat lubricant, wood preservative and Indigenous pesticides, while the remaining articles assessed pure kerosene specifically. Eleven (65%) of these kerosene-specific reports ranked of higher quality (Benítez Riesco et al., 2024; Chattopadhyay et al., 2021; Esashi et al., 2021; Goenka et al., 2022; Gupta et al., 2017; Guss et al., 2020; Hara et al., 2018; Kim et al., 2018; Oreh et al., 2023; Ravikanth et al., 2018; Sanju et al., 2020).

#### 3.4.2. Exposure Setting

Exposure through residential (N=12) and occupational (N=11) settings were most frequently reported across studies; however, exposure contexts varied widely by fuel type. Of the kerosene-specific studies that disclosed an exposure setting, all but one reported instances occurring in the home (N=11), whereas only one study reported residential exposure to jet fuel (Miko et al., 2023). Nearly all jet fuel-specific studies assessed occupational exposure (N=10), predominantly in military (N=9) versus nonmilitary settings (N=2). In contrast, only one study reported workplace exposure to kerosene (N=1). Exposures occurring in the community for both kerosene (N=4) and jet fuel (N=2) were less frequently reported.

#### 3.4.3. Exposure Routes

Patients and participants were most exposed through ingestion (N=14) and inhalation (N=14) across all included studies. Dermal exposures were also commonly reported (N=10). When stratified by fuel type the predominant routes of exposure varied widely; studies assessing kerosene most frequently involved ingestion (N=12, 71%), with fewer documenting inhalation (N=5, 29%) or dermal (N=2, 12%) routes. In contrast, jet fuel was primarily experienced through inhalation (N=9, 82%) and dermal (N=8, 73%) routes. Notably, oral exposure to jet fuel was identified in only two publications (Fuente et al., 2019; Miko et al., 2023).

#### 3.4.4. Exposure Assessment Methods

Exposure was most frequently established through self-reported data (N=24, 86%). This was especially common in case reports, case series, and cross-sectional studies which made up the majority of included studies. All kerosene-specific publications (N=17, 100%), and many jet fuel studies (N=7, 64%), relied on patient or caregiver-reported exposure documented during clinical evaluations. Only three studies, all of which assessed jet fuel, conducted formal exposure assessments: Two analytical studies sampled personal air (Dreisbach et al., 2022; Heaton et al., 2017), and urine (Heaton et al., 2017). In the third study, proxies including task group and job history, self-reported exposure level, and previous industrial assessments, were utilized to estimate exposures (Fuente et al., 2019). One descriptive cross-sectional study identified occupational role (e.g. aircraft ground crew) as a proxy to presume exposure to aviation turbine fuel and lubricants, but did not assess exposure levels or compare outcomes across exposure gradients (Radhakrishnan et al., 2017).

### 3.4 Health Outcomes by Bodily System and Fuel Type

A detailed overview of frequency counts for individual symptom categories within each bodily system, by fuel type and study quality, is provided in Tables 5-6.

**Table 5.**
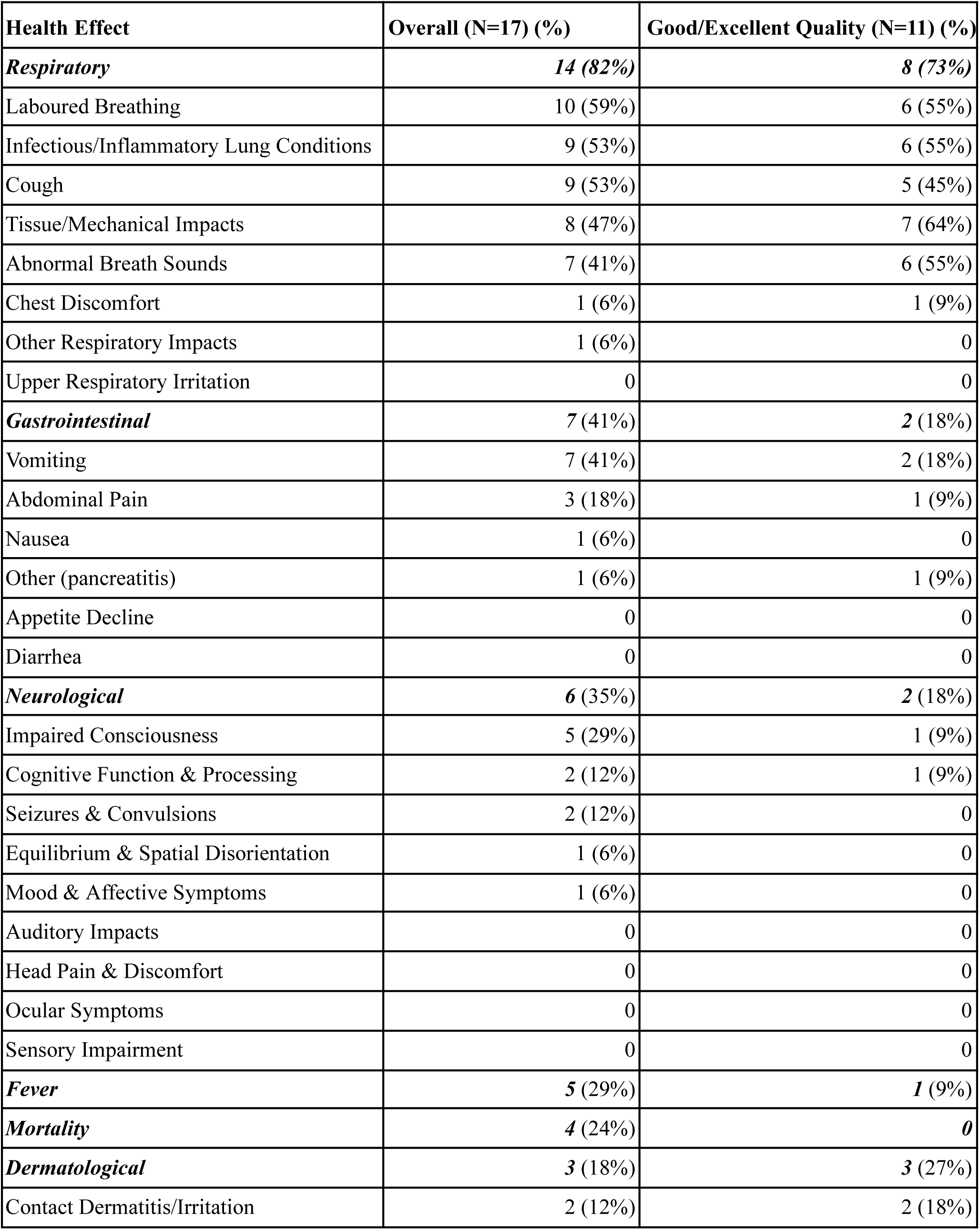
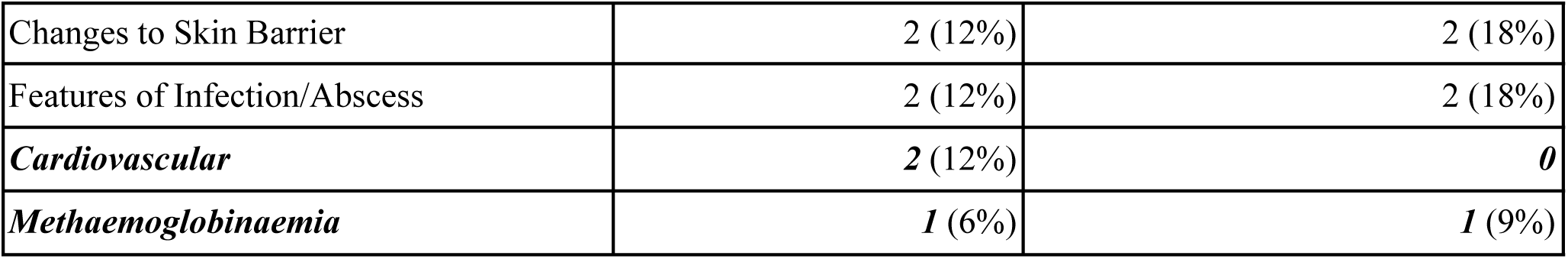
Health Effects Reported Across Kerosene-Specific Studies.

| Health Effect | Overall (N=17) (%) | Good/Excellent Quality (N=11) (%) |
| --- | --- | --- |
| ***Respiratory*** | ***14 (82%)*** | ***8 (73%)*** |
| Laboured Breathing | 10 (59%) | 6 (55%) |
| Infectious/Inflammatory Lung Conditions | 9 (53%) | 6 (55%) |
| Cough | 9 (53%) | 5 (45%) |
| Tissue/Mechanical Impacts | 8 (47%) | 7 (64%) |
| Abnormal Breath Sounds | 7 (41%) | 6 (55%) |
| Chest Discomfort | 1 (6%) | 1 (9%) |
| Other Respiratory Impacts | 1 (6%) | 0 |
| Upper Respiratory Irritation | 0 | 0 |
| ***Gastrointestinal*** | ***7 (41%)*** | ***2 (18%)*** |
| Vomiting | 7 (41%) | 2 (18%) |
| Abdominal Pain | 3 (18%) | 1 (9%) |
| Nausea | 1 (6%) | 0 |
| Other (pancreatitis) | 1 (6%) | 1 (9%) |
| Appetite Decline | 0 | 0 |
| Diarrhea | 0 | 0 |
| ***Neurological*** | ***6 (35%)*** | ***2 (18%)*** |
| Impaired Consciousness | 5 (29%) | 1 (9%) |
| Cognitive Function & Processing | 2 (12%) | 1 (9%) |
| Seizures & Convulsions | 2 (12%) | 0 |
| Equilibrium & Spatial Disorientation | 1 (6%) | 0 |
| Mood & Affective Symptoms | 1 (6%) | 0 |
| Auditory Impacts | 0 | 0 |
| Head Pain & Discomfort | 0 | 0 |
| Ocular Symptoms | 0 | 0 |
| Sensory Impairment | 0 | 0 |
| ***Fever*** | ***5 (29%)*** | ***1 (9%)*** |
| ***Mortality*** | ***4 (24%)*** | ***0*** |
| ***Dermatological*** | ***3 (18%)*** | ***3 (27%)*** |
| Contact Dermatitis/Irritation | 2 (12%) | 2 (18%) |
| Changes to Skin Barrier | 2 (12%) | 2 (18%) |
| Features of Infection/Abscess | 2 (12%) | 2 (18%) |
| **Cardiovascular** | **2 (12%)** | **0** |
| **Methaemoglobinaemia** | **1 (6%)** | **1 (9%)** |

**Table 6.** Health Effects Reported Across Jet Fuel-Specific Studies.

| Health Effect | Overall (N=11) (%) | Good/Excellent Quality (N=6) (%) |
| --- | --- | --- |
| **Neurological** | **8 (73%)** | **4 (67%)** |
| Auditory Impacts | 5 (45%) | 2 (33%) |
| Cognitive Function & Processing | 5 (45%) | 3 (50%) |
| Equilibrium & Spatial Disorientation | 3 (27%) | 1 (17%) |
| Head Pain & Discomfort | 2 (18%) | 0 |
| Impaired Consciousness | 2 (18%) | 0 |
| Mood & Affective symptoms | 2 (18%) | 0 |
| Ocular Symptoms | 2 (18%) | 0 |
| Sensory Impairment | 2 (18%) | 1 (17%) |
| Seizures & Convulsions | 1 (9%) | 0 |
| **Dermatological** | **5 (45%)** | **2 (33%)** |
| Contact Dermatitis/Irritation | 5 (45%) | 2 (33%) |
| Changes to Skin Barrier | 2 (18%) | 1 (17%) |
| Features of Infection/Abscess | 0 | 0 |
| **Respiratory** | **4 (36%)** | **2 (33%)** |
| Cough | 4 (36%) | 2 (33%) |
| Chest Discomfort | 3 (27%) | 1 (17%) |
| Laboured Breathing | 3 (27%) | 1 (17%) |
| Upper Respiratory Irritation | 2 (18%) | 0 |
| Other Respiratory Impacts | 1 (9%) | 1 (17%) |
| Abnormal Breath Sounds | 1 (9%) | 1 (17%) |
| Infectious/Inflammatory Lung Conditions | 0 | 0 |
| Tissue/Mechanical Impacts | 0 | 0 |
| **Gastrointestinal** | **3 (27%)** | **1 (17%)** |
| Abdominal Pain | 2 (18%) | 1 (17%) |
| Appetite Decline | 2 (18%) | 1 (17%) |
| Nausea | 2 (18%) | 1 (17%) |
| Vomiting | 2 (18%) | 1 (17%) |
| Diarrhea | 1 (9%) | 0 |
| Other | 0 | 0 |
| ***Renal*** | ***2*** (18%) | ***1*** (17%) |
| ***Ocular*** | ***2*** (18%) | ***0*** |
| ***Cardiovascular*** | ***1*** (9%) | ***0*** |
| ***Mortality*** | ***1*** (9%) | ***0*** |
| ***Fever*** | ***1*** (9%) | ***1*** (17%) |

#### 3.4.1 Respiratory

Impacts to the respiratory system were most observed, reported in 18 of the 28 included studies (64%), following oral (N=13), inhalation (N=9) and dermal (N=3) exposures which often co-occurred. The most frequently documented respiratory symptoms following kerosene and jet fuel exposures included laboured breathing (e.g. breathlessness, dyspnea and tachypnea) (N=13) (Dayasiri et al., 2017; Esashi et al., 2021; Fife et al., 2018; Goenka et al., 2022; Hara et al., 2018; Kim et al., 2018; Miko et al., 2023; Oba-Daini et al., 2020; Oreh et al., 2023; Parekh & Gupta, 2017; Poon et al., 2019; Ravikanth et al., 2018; Reddy et al., 2020) and cough (N=13) (Areprekumor et al., 2024; Dayasiri et al., 2017; Esashi et al., 2021; Fife et al., 2018; Goenka et al., 2022; Kim et al., 2018; Miko et al., 2023; Oba-Daini et al., 2020; Oreh et al., 2023; Parekh & Gupta, 2017; Poon et al., 2019; Ravikanth et al., 2018; Salam et al., 2020), followed closely by infectious or inflammatory conditions (N=9), tissue or mechanical impacts (N=8) and abnormal breath sounds (N=8). Less frequently reported symptoms included chest discomfort, upper respiratory irritation and other respiratory impacts (i.e. respiratory distress and decline in exercise tolerance).

When restricting analyses to good or excellent quality studies, respiratory symptoms remained the most frequently reported health effects (N=10) (Chattopadhyay et al., 2021; Esashi et al., 2021; Goenka et al., 2022; Kim et al., 2018; Hara et al., 2018; Oreh et al., 2023; Poon et al., 2019; Ravikanth et al., 2018; Salam et al., 2020; Sanju et al., 2020). Similar symptom patterns were observed in this subset: laboured breathing, tissue or mechanical impacts, abnormal breath sounds and cough were most frequently identified (N=7, each), followed closely by infectious or inflammatory lung conditions (N=6). Upper respiratory irritation however, did not appear in studies of higher quality.

##### 3.4.1.1 Kerosene

Respiratory effects were recorded in the greater part of all kerosene-specific studies (82%, N=14) (Areprekumor et al., 2024; Chattopadhyay et al., 2021; Dayasiri et al., 2017; Esashi et al., 2021; Goenka et al., 2022; Hara et al., 2018; Kim et al., 2018; Oba-Daini et al., 2020; Oreh et al., 2023; Parekh & Gupta, 2017; Ravikanth et al., 2018; Reddy et al., 2020; Sanju et al., 2020; Subramanian et al., 2018). Most respiratory complaints involved oral exposures (N=12), with fewer documenting inhalation (N=5). Laboured breathing (N=10) (Dayasiri et al., 2017; Esashi et al., 2021; Goenka et al., 2022; Hara et al., 2018; Kim et al., 2018; Oba-Daini et al., 2020; Oreh et al., 2023; Parekh & Gupta, 2017; Ravikanth et al., 2018; Reddy et al., 2020) and infectious or inflammatory lung conditions, including aspiration pneumonia and chemical pneumonitis (N=9) (Areprekumor et al., 2024; Chattopadhyay et al., 2021; Esashi et al., 2021; Goenka et al., 2022; Hara et al., 2018; Kim et al., 2018; Parekh & Gupta, 2017; Ravikanth et al., 2018; Subramanian et al., 2018), appeared most in kerosene-specific studies. Cough (N=9), tissue or mechanical impacts (N=8), as well as abnormal breath sounds (N=7) were also commonly noted, whereas chest discomfort and general respiratory distress appeared less. Upper respiratory irritation was not reported following kerosene exposure, and no respiratory symptoms were observed with dermal contact.

Among the 13 studies reporting health effects from kerosene exposure, eight were of good or excellent quality (Chattopadhyay et al., 2021; Esashi et al., 2021; Goenka et al., 2022; Hara et al., 2018; Kim et al., 2018; Oreh et al., 2023; Ravikanth et al., 2018; Sanju et al., 2020). Tissue or mechanical impacts such as decreased air entry and emphysematous changes, were the most prevalent respiratory impacts (N=7) within these studies, followed closely by laboured breathing, infectious or inflammatory lung conditions, abnormal breath sounds (N=6, each), and cough (N=5). Chest discomfort, while less prevalent, was also supported by higher quality evidence.

##### 3.4.1.2 Jet Fuel

Among 11 jet-fuel specific studies, respiratory effects were documented in four (36%), all of which involved inhalation of JP-4, JP-5, or JP-8 specifically (Fife et al., 2018; Miko et al., 2023; Poon et al., 2019; Salam et al., 2020) however, dermal (N=3) and oral (N=1) routes were often co-occurring. Cough was the most consistent respiratory complaint, reported in each of the four studies (Fife et al., 2018; Miko et al., 2023; Poon et al., 2019; Salam et al., 2020). Symptoms of chest discomfort such as angina, chest pressure and pleuritic pain, (Fife et al., 2018; Miko et al., 2023; Poon et al., 2019) and laboured breathing (Fife et al., 2018; Miko et al., 2023; Poon et al., 2019) were also detected. Upper respiratory irritation and abnormal breath sounds were less prevalent among these studies, with one documenting a decline in exercise tolerance.

Respiratory symptoms following jet fuel exposure were supported by two studies of higher quality (Poon et al., 2019; Salam et al., 2020) with reports of cough consistent across both. Chest discomfort, laboured breathing, abnormal breath sounds and other respiratory impacts were each reported in only one study of notable (good or excellent) quality. No higher quality evidence suggested infectious or inflammatory lung conditions, tissue or mechanical impacts, or upper respiratory irritation with jet fuel exposure.

#### 3.4.2 Neurological and Auditory

Evidence of neurological and/or auditory impacts were reported in 14 studies (50%) (Areprekumor et al., 2024; Bateganya & Nakanjako, 2023; Dayasiri et al., 2017; Dreisbach et al., 2022; Esashi et al., 2021; Fife et al., 2018; Fuente et al., 2019; Hara et al., 2018; Heaton et al., 2017; Long & Charles, 2018; Miko et al., 2023; Parekh & Gupta, 2017; Reddy et al., 2020; Salam et al., 2020) following inhalation (N=12), oral (N=7), and dermal (N=6) exposures to kerosene and/or jet fuel. Impaired consciousness (Areprekumor et al., 2024; Dayasiri et al., 2017; Fife et al., 2018; Hara et al., 2018; Miko et al., 2023; Parekh & Gupta, 2017; Reddy et al., 2020), and impacts on cognitive function and processing were the most consistent neurological effects overall (N=7, each) (Esashi et al., 2021; Fife et al., 2018; Fuente et al., 2019; Heaton et al., 2017; Miko et al., 2023; Parekh & Gupta, 2017; Salam et al., 2020). These symptoms were followed closely by auditory impacts (N=6) (Dreisbach et al., 2022; Fife et al., 2018; Fuente et al., 2019; Heaton et al., 2017; Long & Charles, 2018; Miko et al., 2023).

When limited to studies of notable quality, neurological outcomes remained the second most frequently reported across included studies (N=6) (Esashi et al., 2021; Fuente et al., 2019; Hara et al., 2018; Heaton et al., 2017; Long & Charles, 2018; Salam et al., 2020). Higher quality evidence supported impacts on cognitive function and processing to be the most consistent neurological symptoms (N=4). Auditory impacts, equilibrium and spatial disorientation, impaired consciousness and sensory impairment were less frequently observed among these reports. Other symptoms observed in lower quality studies such as head pain and discomfort, mood, affective, or ocular symptoms, or seizures and convulsions, were not supported by findings of good or excellent quality reports.

##### 3.4.2.1 Kerosene

Neurological impacts were recorded in 35% of studies reporting on kerosene (N=6) (Areprekumor et al., 2024; Dayasiri et al., 2017; Esashi et al., 2021; Hara et al., 2018; Parekh & Gupta, 2017; Reddy et al., 2020), largely connected to oral (N=5) and inhalation (N=4) exposures. Although impaired consciousness (e.g. delirium, drowsiness and fainting) was most identified (N=5) (Areprekumor et al., 2024; Dayasiri et al., 2017; Hara et al., 2018; Parekh & Gupta, 2017; Reddy et al., 2020), only one report ranked above fair quality (Hara et al., 2018).

Two studies of good or excellent quality noted neurological effects of kerosene exposure (Esashi et al., 2021; Hara et al., 2018), with one report of impaired consciousness, cognitive function and processing. Other symptoms including seizures and convulsions, mood and affective impacts as well as equilibrium and spatial disorientation appeared less and were not supported by higher quality evidence. No studies, regardless of quality, observed auditory impacts or sensory impairment following kerosene exposure.

##### 3.4.2.2 Jet Fuel

Neurological and/or auditory symptoms following jet fuel were identified in eight publications (73%) (Bateganya & Nakanjako, 2023; Dreisbach et al., 2022; Fife et al., 2018; Fuente et al., 2019; Heaton et al., 2017; Long & Charles, 2018; Miko et al., 2023; Salam et al., 2020). While all instances involved inhalation exposure, dermal (N=6) and oral (N=2) exposures were often co-occurring.

Four studies were rated of good or excellent quality (Fuente et al., 2019; Heaton et al., 2017; Long & Charles, 2018; Salam et al., 2020). Impaired cognitive function and processing (including visual memory impacts, confusion, and difficulty concentrating), were among the most widely reported neurological symptoms overall (Fife et al., 2018; Fuente et al., 2019; Heaton et al., 2017; Miko et al., 2023; Salam et al., 2020), and in higher quality studies (Fuente et al., 2019; Heaton et al., 2017; Salam et al., 2020). While indicators of equilibrium and spatial disorientation such as dizziness, stumbling, and sensory impairment (e.g. loss of pain, cold and hunger sensations) were reported less, these symptoms also appeared in at least one study of notable quality. In contrast, head pain and discomfort, impaired consciousness, mood, affective and ocular symptoms, as well as seizures and convulsions were documented less overall and only in poorer quality studies.

Two high quality analytical studies tested associations between jet fuel and neurological outcomes. Both observed higher JP-8 inhalation to be associated with poorer cognitive function and processing among male and female adults of working age (Fuente et al., 2019; Heaton et al., 2017). Fuente et al. reported a dose-response relationship as increased inhalation, dermal and oral exposure resulted in poorer central auditory processing and compressed speech among members of the Royal Australian Air Force (2019). Similarly, greater JP-8 inhalation in a cohort of U.S. Air Force personnel were associated with decreased performances in visual memory and motor speed, though fine motor performance (measured through Grooved Pegboard Test) improved among those more highly exposed (Heaton et al., 2017).

Auditory impacts following jet fuel exposure were documented in five studies (45%), most of which were fair quality (Dreisbach et al., 2022; Fife et al., 2018; Miko et al., 2023), with only two scoring good or excellent (Fuente et al., 2019; Long & Charles, 2018). In one case report of excellent quality, a military aviation technician of 22 years of age complained of severe ear pain, drainage, muffled hearing and a sensation of ear pressure following an incident in which JP-5 entered his ear canal after splashing his face (Long & Charles, 2018). Audiometric testing, however, confirmed the patient’s hearing was within normal limits (Long & Charles, 2018). These findings contrast with those of Fuente et al. who, in an analytical cross sectional study of good quality, observed higher JP-8 exposures among Royal Australian Air Force personnel to be associated with significantly worse hearing thresholds (at 4kHZ in both ears, and 8kHz in the right ear) and poorer average hearing thresholds across all frequencies (1-8kHz) in participant’s “better” ear (2019). Additionally, increased exposure was associated with significantly worse results on pure-tone thresholds, distortion product otoacoustic emissions, auditory brainstem response wave V latency, and scores for words-in-noise (Fuente et al., 2019). With these results, Fuente et al. suggest that jet fuel, when combined with noise exposure, adversely impacts the human auditory system (2019). Similar symptoms were reported among studies of poorer quality, including increased difficulty in listening situations, worse speech intelligibility in noise, ear pain, hearing loss, and ringing in ears, following oral, dermal and inhalation exposures to jet fuel.

#### 3.4.3 Gastrointestinal

Reports of gastrointestinal effects were present in ten studies (36%) (Areprekumor et al., 2024; Bateganya & Nakanjako, 2023; Dayasiri et al., 2017; Miko et al., 2023; Oba-Daini et al., 2020; Oreh et al., 2023; Parekh & Gupta, 2017; Reddy et al., 2020; Salam et al., 2020; Sanju et al., 2020), with vomiting being the most prevalent symptom overall (N=9) (Areprekumor et al., 2024; Dayasiri et al., 2017; Miko et al., 2023; Oreh et al., 2023; Parekh & Gupta, 2017; Reddy et al., 2020; Salam et al., 2020; Sanju et al., 2020). Gastrointestinal symptoms often stemmed from oral (N=8) or inhalation (N=5) exposures, with dermal contact (N=2) noted less among these reports.

Among good or excellent quality studies reporting gastrointestinal effects, vomiting was consistently documented in each (Oreh et al., 2023; Salam et al., 2020; Sanju et al., 2020). Although abdominal pain, decline in appetite, nausea and pancreatitis appeared less overall, these symptoms were supported by at least one higher quality study. In contrast, diarrhea was reported exclusively in one poorer quality report (Miko et al., 2023).

##### 3.4.3.1 Kerosene

Gastrointestinal effects were observed in seven kerosene-specific studies (41%) (Areprekumor et al., 2024; Dayasiri et al., 2017; Oba-Daini et al., 2020; Oreh et al., 2023; Parekh & Gupta, 2017; Reddy et al., 2020; Sanju et al., 2020) often following ingestion (N=7), whereas inhalation was noted in only two of these reports (Areprekumor et al., 2024; Dayasiri et al., 2017). Vomiting was consistently reported in each of these studies, two of which were higher quality (Oreh et al., 2023; Sanju et al., 2020). Abdominal pain and pancreatitis were also observed in one report of higher quality (Sanju et al., 2020), while nausea was documented in one lower quality study (Dayasiri et al., 2017).

##### 3.4.3.2 Jet Fuel

Gastrointestinal symptoms were recorded in 27% (N=3) of jet fuel-specific studies (Bateganya & Nakanjako, 2023; Miko et al., 2023; Salam et al., 2020), all of which followed inhalation exposure, often in combination with dermal (N=2) and oral (N=1) routes. While no symptom was recorded consistently across these studies, vomiting, abdominal pain, appetite decline, and nausea appeared most within this subset (N=2 each). These findings were further supported by one high quality case report (Salam et al., 2020). Diarrhea however, was noted in only one lower quality study (Miko et al., 2023).

#### 3.4.4 Dermatological

Dermatological manifestations were documented in 29% of all included studies (N=8) (Benítez Riesco et al., 2024; Contestable, 2017; Fife et al., 2018; Guss et al., 2020; Hara et al., 2018; Long & Charles, 2018; Miko et al., 2023; Radhakrishnan et al., 2017), nearly all involving dermal contact (N=7). Inhalation (N=4) and oral (N=1) exposures were noted less among these reports. Signs of contact dermatitis and/or irritation such as dry, itchy or burning skin, were the most frequently reported dermatological symptoms overall (N=7) (Benítez Riesco et al., 2024; Contestable, 2017; Fife et al., 2018; Guss et al., 2020; Long & Charles, 2018; Miko et al., 2023; Radhakrishnan et al., 2017), and among higher quality studies (N=4) (Benítez Riesco et al., 2024; Contestable, 2017; Guss et al., 2020; Long & Charles, 2018). Although less frequently observed, features of infection or abscess such as ulcers, purulence and phlegmonous changes, as well as skin barrier changes (e.g. desquamation, whitening of skin) were evident in both higher and lower quality studies.

##### 3.4.4.1 Kerosene

Three case reports, all of good quality, reported dermatological symptoms (18%) following dermal (Benítez Riesco et al., 2024; Guss et al., 2020) and inhalation (Hara et al., 2018) exposures to kerosene. Contact dermatitis and/or irritation (Benítez Riesco et al., 2024; Guss et al., 2020), changes to the skin barrier (Benítez Riesco et al., 2024; Hara et al., 2018) and features of infection and/or abscess (Benítez Riesco et al., 2024; Guss et al., 2020) were equally reported following kerosene exposure. Additional evidence of dermatological effects following kerosene exposure was not demonstrated among poorer quality studies.

##### 3.4.4.2 Jet Fuel

Five jet-fuel specific studies (45%) observed dermatological outcomes, and each noted contact dermatitis and/or irritation following dermal exposure (Contestable, 2017; Fife et al., 2018; Long & Charles, 2018; Miko et al., 2023; Radhakrishnan et al., 2017). Inhalation (N=3) and oral (N=1) exposures were frequently identified alongside dermal contact. Two studies stemmed from higher quality case reports of male U.S. military servicemembers who experienced dermal (Contestable, 2017; Long & Charles, 2018) and inhalation (Long & Charles, 2018) exposures to JP-5. In one case, a 19 year-old U.S. Navy sailor developed a pruritic rash on his hands and forearms that worsened over 10 days following direct contact with a large volume of JP-5 and was ultimately diagnosed with irritant contact dermatitis (Contestable, 2017). While the reported symptoms seemed to follow an isolated incident, the patient reported routine occupational exposure to JP-5 during storing, transferring and refueling operations (Contestable, 2017). Similarly, a 22 year-old military aviation technician was diagnosed with acute non-infectious otitis externa after JP-5 entered his ear canal (Long & Charles., 2018). Clinical evaluation revealed erythema, and skin barrier alterations including a wet-appearance with debris consisting of large amounts of gray to whitish desquamating skin (Long & Charles, 2018).

Changes to skin barrier were less observed, and appeared in both higher and lower quality studies. Unlike kerosene, features of infection or abscess following jet fuel exposure were not observed among included studies.

#### 3.4.5 Less Reported Health Effects

Infrequent health effects linked to kerosene and/or jet fuel exposures included mortality, cardiovascular, ocular, renal, and miscellaneous (i.e. fever and methemoglobinemia) outcomes, each reported in fewer than six studies. Higher-quality studies also identified fever (N=3), methaemoglobinaemia, and renal disease as potential outcomes (Goenka et al., 2022; Gupta et al., 2017; Salam et al., 2020).

##### 3.4.5.1. Mortality

Mortality outcomes following kerosene or jet fuel were documented in five included studies, and exclusively among children (Areprekumor et al., 2024; Bateganya & Nakanjako, 2023; Dayasiri et al., 2017; Parekh & Gupta, 2017; Reddy et al., 2020). Among the four kerosene-specific studies, death followed ingestion in pediatric patients as young as 9 months old (Areprekumor et al., 2024; Dayasiri et al., 2017; Parekh & Gupta, 2017; Reddy et al., 2020), with inhalation documented as a potential co-occurring exposure route in two of these reports (Areprekumor et al., 2024; Dayasiri et al., 2017). Similar findings were identified in a descriptive ethnographic study where premature death was one of two primary side effects of sniffing aviation fuel among child migrants living on the Uganda-Kenya border (Bateganya & Nakanjako, 2023). However, no evidence of higher quality indicated mortality as an outcome of kerosene or jet fuel exposures.

##### 3.4.5.2. Cardiovascular

Cardiovascular symptoms including erratic heart beats, tachycardia and cardiopulmonary arrest, were documented in three studies following kerosene (Dayasiri et al., 2017; Reddy et al., 2020) and jet fuel (Fife et al., 2018) exposures. In both kerosene-specific studies, cardiovascular effects including cardiopulmonary arrest resulting in death, occurred in children aged 9 months to 12 years following ingestion of kerosene or a kerosene-based liquid mosquito repellent. Other cardiovascular symptoms, including palpitations and erratic heart beats, were observed in two female U.S. military personnel with 3-5 years of JP-8 inhalation in a poorly ventilated workplace (Fife et al., 2018). No high quality studies provided evidence supporting cardiovascular effects of kerosene or jet fuel exposure.

##### 3.4.5.3. Ocular

Ocular effects appeared in two publications, both involving jet fuel exposures (Fife et al., 2018; Miko et al., 2023). Symptoms such as eye irritation and burning, often persisting for more than 30 days post exposure, occurred in both children and adults following oral, dermal and inhalation exposure to JP-5 contaminated groundwater (Miko et al., 2023). Similar symptoms were identified in two female U.S. military personnel who presented with blurred vision and eye irritation after 3-5 years of occupational JP-8 and JP-4 exposures (Fife et al., 2018). Both reports however were of lower quality, and no ocular symptoms were present in higher quality studies or those related to kerosene specifically.

##### 3.4.5.4. Renal

Two accounts of renal effects were identified, both in adults of working age following chronic dermal and inhalation exposure to jet fuel (Fife et al., 2018; Salam et al., 2020). One case report of higher quality documented acute renal failure in a 50 year old male aircraft refueler who experienced four years of daily inhalation and dermal exposure to jet fuel (Salam et al., 2020). Renal effects of jet fuel were further supported by one lower quality case report which outlined recurrent bladder and urinary tract infections in two U.S. female service members following chronic JP-8 and JP-4 inhalation (Fife et al., 2018).

##### 3.4.5.5. Other Health Effects

Miscellaneous health effects included reports of fever (N=6) and one hematologic diagnosis (N=1).

Incidences of fever predominantly followed ingestion of kerosene, in both children (Areprekumor et al., 2024; Oba-Daini et al., 2020; Parekh & Gupta, 2017; Reddy et al., 2020), and adults (Goenka et al., 2022). Only one high quality case report documented fever, where an adult male patient presented with acute renal failure following chronic inhalation and dermal exposure to jet fuel (Salam et al., 2020).

Hematological effects were outlined in only one study, in which a 28 year old male was diagnosed with methaemoglobinaemia following ingestion of a kerosene-based Indigenous pesticide in India (Gupta et al., 2017). While no additional hematological effects were identified, laboratory and clinical measurements, including oxygen saturation levels or blood pressure, were not included in this review unless presented as observable symptoms or clinical diagnoses.

Ultimately, evidence of cancer, endocrine, developmental, hepatic, immune, reproductive, musculoskeletal and metabolic outcomes following exposure to pre-combustion forms of kerosene or jet fuel were not identified among included studies.

## 4. Discussion

Our work builds upon the 2017 ATSDR review of jet fuels, with a specific focus on pre-combustion forms of kerosene and kerosene-based jet fuels, which has been absent from previous reviews on the topic. Further, it sought to expand the current knowledge-base on health effects from these fuels by purposely extracting information on diverse population groups and settings, which is essential to understanding potential health effects associated with the widespread environmental contamination from these fuels. This contrasts with much of the previous research which predominantly focused on occupational exposure (U.S. Department of Veterans Affairs, 2023; Vincent-Hall et al., 2025).

This study, which includes 28 publications, finds the respiratory and neurological systems to be most frequently affected overall. Gastrointestinal and dermatological outcomes were also commonly observed, noted in approximately one third of all included studies. However, when stratified by fuel type, respiratory outcomes were most consistently reported following kerosene-specific exposures, whereas neurological effects appeared most among jet fuel specific studies. This review did not observe evidence of cancer, endocrine, developmental, hepatic, immune, reproductive, musculoskeletal or metabolic outcomes following exposure to either fuel type.

### 4.1. Scientific Evidence of Health Outcomes Following Exposure

The most consistent evidence across all studies included respiratory, neurological, gastrointestinal and dermatological outcomes following kerosene and jet fuel exposures. These patterns are further supported when limiting analyses to higher quality studies, where dermatological effects only increase in prevalence among kerosene-specific reports. Although there is clear overlap in the four systems most impacted overall, stratification by fuel type demonstrates differences in relative frequency and presentation of most prominent health effects. These variations may be reflective of differences in chemical composition, as well as duration, dosage and route of exposures.

Respiratory effects appeared in the greater part of all kerosene-specific studies (82%), particularly as infectious or inflammatory lung conditions, laboured breathing, tissue or mechanical impacts, abnormal breath sounds and cough. These findings mirror those consolidated by ATSDR, which documented human respiratory outcomes such as pneumonitis, cough, dyspnea, pulmonary edema and lung infiltrates following kerosene ingestion (2017). In contrast, in jet fuel-specific studies, respiratory effects were evident in approximately one third of reports (36%), with the most common symptoms being cough, chest discomfort and laboured breathing. For these studies, neurological outcomes, notably auditory impacts and impaired cognitive function and processing, were most widely reported (73%), whereas neurological effects were documented less following kerosene exposures (38%), and manifested primarily as impaired consciousness. Gastrointestinal effects such as vomiting, abdominal pain, decline in appetite, nausea and diarrhea, were recorded in roughly one third (36%) of included studies. These symptoms were more often identified within kerosene-specific literature, which may reflect the frequent ingestion-specific context among these reports. This pattern is consistent with prior reports which have documented these outcomes (e.g. vomiting, abdominal pain, gastroenteritis and diarrhea) in children following kerosene ingestion (ATSDR, 2017). Vomiting in particular was the most consistently recorded gastrointestinal symptom in kerosene-specific studies. While evidence of vomiting was also identified following jet fuel exposures, the limited number of studies reporting gastrointestinal effects in this subset (N=3) precludes meaningful comparisons of most prevalent gastrointestinal symptoms between fuel types. Similar challenges arise when comparing dermatological outcomes between fuel types, as reports were limited to three kerosene-specific studies and five jet fuel-specific studies; however, contact dermatitis and irritation remained the most common manifestations following both exposures. Changes to skin barrier, while less reported, were also identified with both fuel types. These symptom profiles have been documented in previous reviews, including reports of itching skin, blisters, rashes on hands, chemical allergy, and scaly and weeping skin in workers with greater occupational JP-8 exposure (Ritchie et al., 2011). Moreover, erythema, eczematous lesions and defatting dermatitis were identified in women who regularly handled kerosene (Ritchie et al., 2011). Similar effects, including edema, erythema, dermatitis, and desquamation, have been observed in rats, mice, rabbits and pigs following dermal application of JP-5, JP-8 and Jet A, with symptom severity varying based on duration and concentration of exposure (ATSDR, 2017).

Within each body of literature (i.e. kerosene- vs jet fuel- specific publications), the predominant routes and duration of exposures remained relatively consistent, but differed largely between them. For instance, all kerosene-specific studies involved acute exposure, primarily through ingestion. In contrast, only one jet fuel publication reported an acute event, while the remainder involved chronic occupational exposures (91%). Among these studies, ingestion was rarely observed (18%), while inhalation (82%) and dermal (73%) routes comprised the vast majority of exposures. It has been previously established that the rate and extent of TPH absorption varies depending on how the compounds enter the body (ATSDR, 1999). Thus, it is possible that while a subset of bodily systems (i.e. respiratory, neurological, gastrointestinal and dermatological) may be more susceptible to these exposures generally, the unique vulnerability and symptom manifestation of each system observed may be influenced in part by the route and duration of exposure.

These patterns may also reflect differences in chemical composition between fuel types. Among kerosene-specific studies, most involved exposures to pure kerosene, though some (29%) involved kerosene-based products such as lubricants or pesticides. Jet fuel studies predominantly reported on JP-5 or JP-8 specifically (73%), which contain approximately 99.5% kerosene along with special additives (Kuppusamy et al., 2020). Differing formulations further complicate comparisons between fuel types. Additionally, attributing observed health effects to raw, unburned fuel forms alone is not certain. Kerosene exposures largely involved acute ingestion in low- and middle-income countries where kerosene is often used as household fuel. Similarly, jet fuel exposures primarily occurred in occupational settings, with aviation workers experiencing recurrent exposure over several years. In both settings, it is likely that individuals were exposed to post-combustion byproducts, and other co-occurring environmental pollutants; however, the descriptive nature of most included publications limits the consideration of these potential confounders. For these reasons, we cannot attribute the health effects observed in this review to pre-combustion forms of kerosene and kerosene-based jet fuel exclusively. Despite these limitations, the consistent presence of kerosene as a base compound and the descriptive nature of most publications together highlight key similarities across studies, supporting the potential for shared toxicological patterns such as respiratory, neurological, gastrointestinal and dermatological health implications. Lastly, other system-level health effects were rarely documented within either subset. Thus, the system level alignment, including those most frequently affected, as well as those absent, suggests that respiratory, neurological, gastrointestinal and dermatological systems may be particularly vulnerable to these exposures, and should be assessed further in future studies.

### 4.2. Future Studies Assessing Risk of Kerosene and Jet fuel on Human Health

Very few included studies were analytical in design. Of the three that compared outcomes by exposure level (two of notable quality), all focused exclusively on jet fuel (Fuente et al., 2019; Heaton et al., 2017). The authors observed increasing JP-8 exposure to be associated with decreased neurocognitive performance (e.g., visual memory and motor speed), poorer central auditory processing, and worse hearing thresholds (Fuente et al., 2019; Heaton et al., 2017). Descriptive evidence from jet fuel-specific literature included in this review further support neurological risks, particularly through reports of auditory impacts, and impaired cognitive function and processing (73%). Several similar effects have also been observed in animals exposed to JP-8, including lethargy, ototoxic hearing loss, central auditory dysfunction, and impaired learning in rats (ATSDR, 1999, 2017). Comparable neurological outcomes in humans have been documented in prior reviews as well, including nervous system depression, lethargy, coma, drowsiness, convulsions, restlessness and irritability following kerosene ingestion (ATSDR, 1999, 2017). Together, neurotoxicity appears consistent across analytical and descriptive evidence identified in this review, as well as previous literature on kerosene ingestion, all of which is supported further by findings from animal studies. Efforts should be made to continue exploring the unique risks of jet fuel and kerosene on neurocognitive health. This is of particular importance for exposure during fetal development and early childhood.

Although jet fuel-specific reports typically involved more chronic exposures, their descriptive nature or cross-sectional designs largely prevented the identification of health outcomes requiring longer latency periods. Similarly, kerosene studies often reported on clinical evaluations immediately following poisoning. In the sole cohort study identified in this review, the follow-up period spanned only one work week in order to establish neurological outcomes associated with jet fuel exposures. As a result, the lack of evidence demonstrating cancer, endocrine, developmental, hepatic, immune, reproductive, musculoskeletal and metabolic outcomes is likely indicative of a limited follow up needed to observe these more latent health outcomes, rather than an absence of risk. Future research should prioritize longitudinal designs to better detect longer-latency health effects that may remain undiscovered in short-term studies.

The need to address this research gap is emphasized by the adverse health outcomes undetected by the current review being previously observed in animal studies. These findings have been summarized in the toxicological profile of jet fuel, which this review builds upon (ATSDR, 2017), including cancerous outcomes such as malignant lymphomas and skin cancer following dermal JP-5 exposure, as well as tumours of the uterus and vagina following kerosene ingestion (ATSDR, 1999, 2017). Reproductive and developmental effects resulting from ingestion and inhalation of JP-8 have also been found in mice, including increased maternal deaths, decreased sperm motility, reduced fetal weight, decreased litter size, and suppressed immune function in pups (ATSDR, 2017, Ritchie et al., 2011). Furthermore, decreased body weight - particularly in male, fetal and pregnant rats-appeared in several animal studies following oral and inhalation exposures to both JP-5 and JP-8, but was not identified in this review (ATSDR, 1999, 2017).

Self-reported data informed most included reports, with only three studies conducting formal exposure assessment. Considering that jet fuel and kerosene share complex hydrocarbon mixtures with many other petroleum products, no single compound can indicate their exclusive presence. Although compounds such as naphthalene or benzene can be measured to inform exposure levels, they are widely present in the environment and cannot be used to distinguish contact specifically to pre- versus post- combustion fuel forms (ATSDR, 2017). Therefore, future studies should strive to employ a combination of personal sampling, and contextual factors that demonstrate contact with raw fuel specifically. Similarly, the greater part of all health effects identified by this review stem from descriptive accounts of clinical evaluations which did not account for confounding variables. Given that most jet fuel-specific literature involved occupational settings, and kerosene-specific literature originated primarily from low- and middle- income countries, additional environmental exposures may be unaccounted for. Future studies should address potential confounding or mediating influences of co-occurring pollutants.

The predominant occupational focus among the few analytical studies identified by this review may be due to the recognition of jet fuel as one of the largest chemical exposures facing both military and civilian aviation workers (Ritchie et al., 2003). However, only three studies assessing the association between pre-combustion jet fuel on human health have been identified across the review period (2017-2024). Despite the recognized occupational burden of jet fuel across military and non-military settings alike, regulatory exposure thresholds continue to rely heavily on animal models and *in vitro* toxicological studies. There remains a need for further occupational research to ensure regulatory limits are grounded in accurate and real-world human data. However, recent jet-fuel contamination events, such as the spill at Red Hill on Oʻahu, Hawaiʻi, and in Bucks County, Pennsylvania, highlight the importance of expanding research beyond occupational settings. Future studies should examine the particular risks of non-occupational, and ingestion related exposures as well, as they remain largely unexplored in current research. Efforts should be made to prioritize inclusion of women, children and older adults for whom the evidence is particularly limited.

### 4.3. Study Strengths and Limitations

To our knowledge, this is the first systematic review to evaluate the evidence of known and potential health effects of exposure to pre-combustion forms of kerosene and kerosene-based jet fuels specifically. The utilization of multiple databases as well as the participation of three reviewers in article selection, data extraction and quality assessment, decrease the likelihood and influence of bias. While limiting inclusion to English, French, Dutch, Spanish or Portuguese publications may have omitted relevant evidence, only one study was excluded during full text screening because of language. Additionally, the evaluation of two chemically similar but distinct fuels expanded our evidence base, and enabled observation of a broader range of reported health effects, as well as consistencies, or lack thereof, in symptom patterns between fuel types. As jet fuel research primarily involves occupational dermal and inhalation exposures among men of working age, limiting the review to this subset of literature would likely omit a toxicological understanding of additional exposure contexts, particularly related to raw, unburned fuel. With kerosene comprising the vast majority of jet fuel’s composition, its inclusion in this review allows for an evaluation of potential health effects stemming from ingestion, non-occupational settings and among women, children and older adults. Thus, the wide scope of this review supports a more comprehensive understanding of potential human health outcomes across generally understudied settings and populations in this research area - particularly those exposures occurring in the home and community, and across the lifespan.

Comparing findings among and between groups by age, sex and setting proved difficult given the heterogeneity of fuel type, exposure route and outcomes, as well as the limited number of studies available within each subgroup. While these differences complicate meaningful comparisons across all included studies, consistency in exposure route, setting and age group within each subset of publications (i.e. kerosene- vs jet fuel- specific studies) allowed for natural comparisons, a particularly interesting feature of this review. It is important to note however, that with only 28 included studies - fewer of notable quality - the evidence base is limited, reducing confidence in the patterns observed. Being that the greater part of all included literature consisted of descriptive accounts of health effects observed following exposures (e.g., case reports and case series), and formal testing of associations between these variables was rarely conducted, the ability to establish fuel attributable causality from the findings of this review is not possible.

Given these constraints, a meta-analysis could not be conducted. Finally, the vast differences in exposure duration between fuel-specific studies provide an insight as to potential health effects related to acute versus chronic exposures. These conclusions however, are challenged by key differences in geographical setting, participant age, exposure routes, dosage levels, as well as the chemical composition of each fuel type. Since reports of kerosene poisoning comprised a larger portion of included reports, the data identified, and subsequent findings, may be moderately weighed in favour of acute symptoms. The urgency to explore health implications of chronic exposure is underscored by the scarcity of longitudinal studies, and lack of health effects typically requiring longer latency periods (e.g., reproductive, developmental, and cancerous outcomes), identified in this systematic review.

## 5. Conclusion

The widespread burden of jet fuel in occupational settings and kerosene as household fuel in low- and middle- income countries, as well as recent drinking water contamination events that extend beyond the scope of existing research, underscore the need for further evaluation of potential human health impacts of these fuels. Considering the vast consumption of kerosene and kerosene-based jet fuels globally, widespread anthropocentric releases of TPHs to the environment, and the limitations of current research, efforts should be made to better understand the health implications of exposure to these raw fuels. The importance of this research is supported by this review, which finds consistent evidence of adverse human health outcomes in various settings and across the lifespan. To this end, it is our hope that as evidence is generated through future epidemiological studies, we may be better equipped to evaluate and discern the cumulative impact of exposure to unburned jet fuel and kerosene on human health outcomes.

## Data Availability

All data produced in the present work are contained in the manuscript

## Acknowledgements

We would like to express our sincere gratitude to Carolyn Ching Dennison, Science and Technology Librarian with University of Hawai’i at Mānoa for her contributions to the development of the search strategy utilized in this review.

## Abbreviations

ATSDR: Agency for Toxic Substances and Disease Registry
B/D: Barrels per Day
EIA: Energy Information Administration
EPA: Environmental Protection Agency
JP: Jet Propellant
NIOSH: National Institute for Occupational Safety and Health
PECO: Population, Exposure, Comparator, Outcome
PH: Petroleum hydrocarbons
PRISMA: Preferred Reporting Items for Systematic reviews and Meta-Analyses
PROSPERO: The International Prospective Register of Systematic Reviews
TPH: Total Petroleum Hydrocarbon

---

**Table S1.** Terms Screened and Excluded from Search Strategy*.

| Database | Search terms |
| --- | --- |
| PubMed | "acute effects", angina, anxiety, asthma, blisters, "blurr* vision", body weight, bronchitis, bronchoconstriction, cancer, cardio*, cancerous, cardiovascular, chest tightness, "chronic obstructive pulmonary disease", "chronic illness", coenzymes, congenital abnormalities, congenital diseases, connective tissue, confusion, convulsions, dermatitis, developmental, developmental disorder, diarrhea, digestive, dyspnea, endocrine, emphysema, estrogen, "eye disease", faint*, "heart disease", "hemic diseases", hemorrhage, "hereditary diseases", "hereditary abnormalities", histopatholog*, hormonal, "immunotoxicity", imbalance, inflammation, irrita*, liver injury, lightheaded*, lymph, lymphatic diseases, mental disorder, "mental health", mutation, musculoskeletal, nausea, neoplasm, "neonatal diseases", "neonatal abnormalities", "nervous system", neurological, "ocular", ototoxic, pulmonary lesions, pregnancy complications, pulmonary, quality of life, remember*, reproductive, respiratory, seizures, skin, stomatognathic, systemic effects, tachycardia, tremor, "toxicology", "toxic effects", "toxicological effects", urogenital, "urogenital diseases", "vomit*", "walk*", wheez* |
| Web of Science | "acute effects", angina, asthma, anxiety, bronchitis, blisters, "blurr* vision", "body weight", bronchoconstriction, cardiovascular, "connective tissue", cancerous, cardio*, "chest tight*", cough*, "congenital abnormalities", "congenital diseases", coenzymes, cerebrovascular, "chest tightness", "chronic obstructive pulmonary disease", convulsions, "chronic disease", "chronic effects", "chronic illness", "developmental disorder", dental, developmental, derma*, dyspnea, dizz*, etiolog*, endocrin*, eyes, "eye disease", "eye diseases", emphysema, faint*, forget*, gastro*, gastrointestinal, gestation, genomics, hear*, hepatic, hemorrhage, histopatholog*, "hemic diseases", "hereditary diseases", "hereditary abnormalities", "health outcome", itch*, "immune system", kidney*, "lymphatic diseases", "liver injury", neoplasm, "neonatal diseases", "neonatal abnormalities", musculoskeletal, "mental illness", morbidit*, "mental health", "mental disorder", mutagen, mutation, neurolog*, "nervous system", ocular, "occupational accidents", "out of breath", patholog*, pulmonary, "pregnancy complications", remember*, reproductive, rash, sex, "stomatognathic diseases", symptom, "toxic* effects", urogenital diseases, vision, vertigo, wheez*, worry |

\*Terms not included in search strategy after failing to pull additional results during database search

**Table S2.** Quality Assessment Framework for Cohort & Case-Control Studies.

Max Score: 24
| Item | Criteria | Answers |
| --- | --- | --- |
| **Reporting** |  |  |
| 1 | *Is the hypothesis/aim/objective of the study clearly described?* | Yes = 1<br>No = 0 |
| 2 | *Are the main outcomes to be measured clearly described in the Introduction or Methods section?*<br>If the main outcomes are first mentioned in the Results section, the question should be answered no. | Yes = 1<br>No = 0 |
| 3 | *Are the characteristics of the participants included in the study clearly described?*<br>In cohort studies, inclusion and/or exclusion criteria should be given. In case-control studies, a case-definition and the source for controls should be given. | Yes = 1<br>No = 0 |
| 4 | *Are the distributions of principal confounders in each group of subjects to be compared clearly described?*<br>A list of principal confounders is provided.<sup>a</sup> | Yes = 2<br>Partially = 1<br>No = 0 |
| 5 | *Are the main findings of the study clearly described?*<br>Simple outcome data (including denominators and numerators) should be reported for all major findings so that the reader can check the major analyses and conclusions. (This question does not cover statistical tests which are considered below). | Yes = 1<br>No = 0 |
| 6 | *Does the study provide estimates of the random variability in the data for the main outcomes?*<br>In non-normally distributed data the interquartile range of results should be reported. In normally distributed data the standard error, standard deviation or confidence intervals should be reported. If the distribution of the data is not described, it must be assumed that the estimates | Yes = 1<br>No = 0 |
|  | used were appropriate and the question should be answered yes.<sup>b</sup> |  |
| 7 | *Have the characteristics of participants lost to follow-up been described?*<br>This should be answered yes where there were no losses to follow-up or where losses to follow-up were so small that findings would be unaffected by their inclusion. This should be answered 'no' where a study does not report the number of participants lost to follow-up. | Yes = 1<br>No = 0<br>Unable to determine = 0 |
| 8 | *Have actual probability values been reported (e.g. 0.035 rather than <0.05) for the main outcomes except where the probability value is less than 0.001?<sup>c</sup>* | Yes = 1<br>No = 0 |
| **External validity** |  |  |
| 9 | *Were the subjects asked to participate in the study representative of the entire population from which they were recruited?*<br>The study must identify the source population for participants and describe how they were selected. Participants would be representative if they comprised the entire source population, an unselected sample of consecutive participants, or a random sample. Random sampling is only feasible where a list of all members of the relevant population exists. Where a study does not report the proportion of the source population from which the participants are derived, the question should be answered as unable to determine. | Yes = 1<br>No = 0<br>Unable to determine = 0 |
| 10 | *Were those subjects who were prepared to participate representative of the entire population from which they were recruited?*<br>The proportion of those asked who agreed should be stated. Validation that the sample was representative would include demonstrating that the distribution of the main confounding factors was the same in the study sample and the source population. | Yes = 1<br>No = 0<br>Unable to determine = 0 |
| **Internal validity - bias** |  |  |
| 11 | *If any of the results of the study were based on “data dredging”, was this made clear?*<br>Any analyses that had not been planned at the outset of the study should be clearly indicated. If no retrospective unplanned subgroup analyses were reported, then answer yes. | Yes = 1<br>No = 0<br>Unable to determine = 0 |
| 12 | *In cohort studies, do the analyses adjust for different lengths of follow-up of participants, or in case-control studies, is the time period between the intervention and outcome the same for cases and controls?*<br>Where follow-up was the same for all study participants the answer should be yes. If different lengths of follow-up were adjusted for by, for example, survival analysis the answer should be yes. Studies where differences in follow-up are ignored should be answered no. | Yes = 1<br>No = 0<br>Unable to determine = 0 |
| 13 | *Were the statistical tests used to assess the main outcomes appropriate?*<br>The statistical techniques used must be appropriate to the data. For example nonparametric methods should be used for small sample sizes. Where little statistical analysis has been undertaken but where there is no evidence of bias, the question should be answered yes. If the distribution of the data (normal or not) is not described it must be assumed that the estimates used were appropriate and the question should be answered yes. | Yes = 1<br>No = 0<br>Unable to determine = 0 |
| 14 | *Were the main outcome measures used accurate (valid and reliable)?*<br>For studies where the outcome measures are clearly described, the question should be answered yes. For studies which refer to other work or that demonstrates the outcome measures are accurate, the question should be answered as yes. | Yes = 1<br>No = 0<br>Unable to determine = 0 |
| **Internal validity – confounding (selection bias)** |  |  |
| 15 | *Were the participants in different intervention groups (cohort studies) or were the cases and controls (case-control studies) recruited from the same population?*<br>For example, participants for all comparison groups should be selected from the same hospital. The question should be answered unable to determine for cohort and case-control studies where there is no information concerning the source of participants included in the study.<sup>d</sup> | Yes = 1<br>No = 0<br>Unable to determine = 0 |
| 16 | *Were study subjects in different intervention groups (cohort studies) or were the cases and controls (case-control studies) recruited over the same period of time?* For a study which does not specify the time period over which participants were recruited, the question should be answered as unable to determine.<sup>d</sup> | Yes = 1<br>No = 0<br>Unable to determine = 0 |
| 17 | *Was there adequate adjustment for confounding in the analyses from which the main findings were drawn?*<br>In non-randomized studies if the effect of the main confounders was not investigated or confounding was demonstrated but no adjustment was made in the final analyses the question should be answered as no.<sup>d</sup> | Yes = 1<br>No = 0<br>Unable to determine = 0 |
| 18 | *Were losses of participants to follow-up taken into account?*<br>If the numbers of participants lost to follow-up are not reported, the question should be answered as unable to determine. If the proportion lost to follow-up was too small to affect the main findings, the question should be answered yes. | Yes = 1<br>No = 0<br>Unable to determine = 0 |
| **Power** |  |  |
| 19* | *Was a power analysis performed?*<sup>d</sup> | Yes = 1<br>No = 0<br>Unable to determine = 0 |
| **Exposure Assessment** |  |  |
| 20 | *Was the exposure of interest the focus of the study?*<br>If the exposure was the main variable tested, this question should be answered yes. If the exposure of interest was one of many variables tested, answer no. | Yes = 1<br>No = 0 |
| 21 | *Were the exposure assessment methods used objective and precise?*<br>If the study utilized direct methods to measure exposure levels such as biomonitoring or personal samplers, this question should be answered yes. If the study utilized indirect methods such as questionnaires or self-reported data to infer exposure levels, this question should be answered partially. If the study relied on proxies such as job titles or time spent near exposure source to attribute exposure levels, answer no. | Yes = 2<br>Partially = 1<br>No = 0 |
| 22 | *Is exposure assessed in a way that allows for dose-response conclusions?*<br>If the study assessed outcomes of exposure at several levels of varying intensities or concentrations (ex. low, medium and high exposure), answer yes.<br>If the study categorized exposure into broad groups like “high” and “low” without precise measurements or without including dose-response information, the question should be answered no. | Yes = 1<br>No = 0 |

\*Modified from the original form published by Downs and Black & Carvajal et al., 2025

Quality Assessment Framework for Cross-Sectional Studies Max Score: 21
| Item | Criteria | Answers |
| --- | --- | --- |
| **Reporting** |  |  |
| 1 | *Is the hypothesis/aim/objective of the study clearly described?* | Yes = 1<br>No = 0 |
| 2 | *Are the main outcomes to be measured clearly described in the Introduction or Methods section?*<br>If the main outcomes are first mentioned in the Results section, the question should be answered no. | Yes = 1<br>No = 0 |
| 3 | *Are the characteristics of the participants included in the study clearly described?*<br>Characteristics of participants are included. | Yes = 1<br>No = 0 |
| 4 | *Are the distributions of principal confounders in each group of subjects to be compared clearly described?*<br>A list of principal confounders is provided.<sup>a</sup> | Yes = 2<br>Partially = 1<br>No = 0 |
| 5 | *Are the main findings of the study clearly described?*<br>Simple outcome data (including denominators and numerators) should be reported for all major findings so that the reader can check the major analyses and conclusions. (This question does not cover statistical tests which are considered below). | Yes = 1<br>No = 0 |
| 6 | *Does the study provide estimates of the random variability in the data for the main outcomes?*<br>In non-normally distributed data the interquartile range of results should be reported. In normally distributed data the standard error, standard deviation or confidence intervals should be reported. If the distribution of the data is not described, it must be assumed that the estimates used were appropriate and the question should be answered yes.<sup>b</sup> | Yes = 1<br>No = 0 |
| 7 | *Have actual probability values been reported (e.g. 0.035 rather than $<0.05$) for the main outcomes except where the probability value is less than 0.001?<sup>c</sup>* | Yes = 1<br>No = 0 |
| **External validity** |  |  |
| 8 | *Were the subjects asked to participate in the study representative of the entire population from which they were recruited?*<br>The study must identify the source population for participants and describe how they were selected. Participants would be representative if they comprised the entire source population, an unselected sample of consecutive participants, or a random sample. Random sampling is only feasible where a list of all members of the relevant population exists. Where a study does not report the proportion of the source population from which the participants are derived, the question should be answered as unable to determine. | Yes = 1<br>No = 0<br>Unable to determine = 0 |
| 9 | *Were those subjects who were prepared to participate representative of the entire population from which they were recruited?*<br>The proportion of those asked who agreed should be stated. Validation that the sample was representative would include demonstrating that the distribution of the main confounding factors was the same in the study sample and the source population. | Yes = 1<br>No = 0<br>Unable to determine = 0 |
| **Internal validity - bias** |  |  |
| 10 | *If any of the results of the study were based on “data dredging”, was this made clear?*<br>Any analyses that had not been planned at the outset of the study should be clearly indicated. If no retrospective unplanned subgroup analyses were reported, then answer yes. | Yes = 1<br>No = 0<br>Unable to determine = 0 |
| 11 | *Were the statistical tests used to assess the main outcomes appropriate?*<br>The statistical techniques used must be appropriate to the data. For example nonparametric methods should be used for small sample sizes. Where little statistical analysis has been undertaken but where there is no evidence of bias, the question should be answered yes. If the distribution of the data (normal or not) is not described it must be assumed that the estimates used were appropriate and the question should be answered yes. | Yes = 1<br>No = 0<br>Unable to determine = 0 |
| 12 | *Were the main outcome measures used accurate (valid and reliable)?*<br>For studies where the outcome measures are clearly described, the question should be answered yes. For studies which refer to other work or that demonstrates the outcome measures are accurate, the question should be answered as yes. | Yes = 1<br>No = 0<br>Unable to determine = 0 |
| **Internal validity – confounding (selection bias)** |  |  |
| 13 | *Were the participants recruited from the same population?*<br>This question should be answered yes when the sample was drawn from the same source population. In multi-site studies, answer yes when participants are recruited from similar source populations across all sites. If participant characteristics are not provided, this question should be answered, unable to determine. | Yes = 1<br>No = 0<br>Unable to determine = 0 |
| 14 | *Were study subjects recruited over the same period of time?* For a study which does not specify the time period over which participants were recruited, the question should be answered as unable to determine.<sup>d</sup> | Yes = 1<br>No = 0<br>Unable to determine = 0 |
| 15 | *Was there adequate adjustment for confounding in the analyses from which the main findings were drawn?*<br>In non-randomized studies if the effect of the main confounders was not investigated or confounding was demonstrated but no adjustment was made in the final analyses the question should be answered as no.<sup>d</sup> | Yes = 1<br>No = 0<br>Unable to determine = 0 |
| **Power** |  |  |
| 16* | *Was a power analysis performed?*<sup>d</sup> | Yes = 1<br>No = 0<br>Unable to determine = 0 |
| **Exposure Assessment** |  |  |
| 17 | *Was the exposure of interest the focus of the study?*<br>If the exposure was the main variable tested, this question should be answered yes. If the exposure of interest was one of many variables tested, answer no. | Yes = 1<br>No = 0 |
| 18 | *Were the exposure assessment methods used objective and precise?*<br>If the study utilized direct methods to measure exposure levels such as biomonitoring or personal samplers, this question should be answered yes. If the study utilized indirect methods such as questionnaires or self-reported data to infer exposure levels, this question should be answered partially. If the study relied on proxies such as job titles or time spent near exposure source to attribute exposure levels, answer no. | Yes = 2<br>Partially = 1<br>No = 0 |
| 19 | *Is exposure assessed in a way that allows for dose-response conclusions?*<br>If the study assessed outcomes of exposure at several levels of varying intensities or concentrations (ex. low, medium and high exposure), answer yes. | Yes = 1<br>No = 0 |
|  | If the study categorized exposure into broad groups like “high” and “low” without precise measurements or without including dose-response information, the question should be answered no. |  |

\*Modified from the original form published by Downs and Black & Carvajal et al., 2025

Case Reports & Case Series Critical Appraisal Tool Adapted for Quality Assessment Max Score = 10
| Item | Criteria | Answers |
| --- | --- | --- |
| **Reporting** |  |  |
| 1 | *Were the patient's demographic characteristics clearly described?*<br>Does the case report clearly describe the patient's age, sex, race, medical history, diagnosis and prognosis? The setting and context may also be described. | Yes = 2<br>Some described = 1<br>No = 0 |
| 2 | *Was the presenting clinical condition of the patient on presentation clearly described?*<br>The presenting clinical condition of the patient should be described in detail including the uniqueness of the condition/disease, symptoms, frequency and severity. The case report should also be able to present whether differential diagnoses was considered. | Yes = 2<br>Some described = 1<br>No = 0 |
| 3 | *Were diagnostic tests or assessment methods and the results clearly described?*<br>A reader of the case report should be provided sufficient information to understand how the patient was assessed. The case report should provide a clear description of various diagnostic tests used. | Yes = 1<br>No = 0 |
| 4 | *Was the intervention(s) or treatment procedure(s) clearly described?*<br>The report should describe the treatment/intervention protocol in detail; for e.g. in pharmacological management of dental anxiety - the type of drug, route of administration, drug dosage and frequency, and any side effects. | Yes = 2<br>Some described = 1<br>No = 0 |
| 5 | *Was the post-intervention clinical condition clearly described?*<br>The case report should clearly describe the clinical condition post-intervention in terms of the presence or lack thereof symptoms. | Yes = 1<br>No = 0 |
| 6 | *Were adverse events (harms) or unanticipated events identified and described?*<br>Any adverse events after administering treatment/intervention/drug are clearly documented and described. If authors mention no adverse events occurred after treatment/intervention, then answer yes. | Yes = 1<br>No = 0 |
| 7 | *Does the case report provide takeaway lessons?*<br>Case reports should summarize key lessons learned from a case in terms of the background of the condition/disease and clinical practice guidance for clinicians when presented with similar cases. | Yes = 1<br>No = 0 |

Modified from: Moola S, Munn Z, Tufanaru C, Aromataris E, Sears K, Sfetcu R, Currie M, Qureshi R, Mattis P, Lisy K, Mu P-F. Chapter 7: Systematic reviews of etiology and risk. In: Aromataris E, Munn Z (Editors). Joanna Briggs Institute Reviewer's Manual. The Joanna Briggs Institute, 2017. Available from https://reviewersmanual.joannabriggs.org/

**Table S3.** Categorization of Extracted Health Effects by Bodily System*.

| **Bodily System** | **Symptom Category** | **Raw Symptoms Grouped** |
| --- | --- | --- |
| **Respiratory** | Abnormal Breath Sounds | Bibasal coarse crackles; bilateral coarse crackles; breath sounds decrease with inspiratory rales and crackle on right side; crepitations; decreased breath sounds; decreased breath sounds over right hemithorax; diminished breath sounds; fine crepitations heard in middle and lower lungs; inspiratory rales and crackles bilaterally; noisy breathing; transmitted sounds |
|  | Chest Discomfort | Angina; burning lungs; chest pressure; chest tightness/pain/angina; pleuritic pain; sharp right sided and pleuritic chest pain |
|  | Cough | Cough; dry cough |
|  | Infectious/Inflammatory Lung Conditions | Aspiration pneumonia; chemical pneumonitis; pneumonitis; pneumonia; lipid pneumonia |
|  | Laboured Breathing | Breathlessness; bronchial breathing; dyspnea; laboured breathing; shortness of breath; tachypnea; tachypneic; tachypnoea; rapid breathing; wheeze |
|  | Tissue/Mechanical Impacts | Bilateral dependent dense consolidation and multiple pneumatoceles (some showing air fluid levels); bilateral infiltrates; bronchovascular markings; bronchopleural Fistula (BPF); burn injuries in respiratory tract; consolidation in bilateral lower lobes; costophrenic angle blunting with thickening of right minor fissure; decreased air entry to right lung; double gastric fluid level and consolidation in right middle and left lower lobes; edematous and erythematous changes in right middle and lower lobe with grayish secretions; emphysematous changes; empyema; right middle lobe/right lower lobe consolidation; ill-defined areas of ground-glass opacities with superimposed interlobular septal thickening (crazy-paving pattern) with predominantly lower lobar distribution and low attenuating (lipid-containing) opacities; increased bilateral hilar shadow; infiltration in both lower lobes with low-density area in infiltrative shadow; loculated collection in right middle and lower lobe with pleural thickening; mild right sided pleural effusion; necrotic consolidation; ground glass opacity (GGO) and bronchial wall thickening in right middle/lower lobe; patchy air space consolidation in right lower zone of lung; |
|  |  | patchy infiltrates in bilateral lower zones; pleural collection along right posterior chest wall with air fluid level and posterior basal subsegmental bronchus leading to collection; surrounding subsegmental regional consolidation with mild contralateral mediastinal shift; pleural effusion on right side with collapse; pneumothorax; pus pockets found in middle and lower lobe of right lung right infiltrates; right pleural effusion; thick pus; volume loss with air-fluid level in retrocardiac region on right side |
|  | Upper Respiratory Irritation | Burning nose or throat; nose bleeds; runny nose; sinus congestion; sinus infections; sneezing; upper respiratory infection |
|  | Other Respiratory Impacts | Acute respiratory distress syndrome; decline in exercise tolerance; respiratory distress |
| **Neurological** | Auditory Impacts | Changes to audiometric thresholds; difficulty in listening situations; ear pain; ear pressure; lower speech intelligibility in noise; muffled hearing; ringing in ears; worse hearing thresholds |
|  | Cognitive Function & Processing | Confusion; confusion; difficulty concentrating; difficulty remembering; mild right frontal hypoperfusion; performance with non-dominant hand on Grooved pegboard task; poor central auditory processing (performances in visual memory and motor speed); visual memory |
|  | Equilibrium & Spatial Disorientation | Bilateral vestibular dysfunction on rotational chair test; dizziness; dizzy; fall on Romberg test; feeling of drifting to one side of hallway, overcorrecting and stumbling; general impairment of equilibrium and predominant vestibular deficit pattern; giddiness; imbalance; impaired equilibrium; persistent dizziness; reduced caloric vestibular responses; reduced gain with sinusoidal rotation; sensation of spinning; sensation of tilting; worsening dizziness |
|  | Head Pain & Discomfort | Headache; head fullness |
|  | Impaired Consciousness | Altered sensorium (irritability, lethargy, drowsiness); delirium; drowsiness; fainting; fatigue; loss of consciousness; stupor; unusual sleepiness |
|  | Mood & Affective Symptoms | Agitation/irritability; anxiety; depression; difficulty sleeping; feeling depressed; |
|  |  | paranoia; restlessness; tension/nervousness |
|  | Seizures & Convulsions | Convulsions; seizures |
|  | Sensory Impairment | Feeling numb; loss of pain, cold, hunger sensations; numbness |
| **Gastrointestinal** | Abdominal Pain | Abdominal pain; epigastric pain |
|  | Appetite Decline | Appetite decline; decline in appetite/oral intake; loss of appetite |
|  | Diarrhea | Diarrhea |
|  | Nausea | Nausea |
|  | Vomiting | Vomiting |
|  | Other | Pancreatitis |
| **Dermatological** | Contact Dermatitis/Irritation | Cracking; crusting; dry or itchy skin; edema; erythema; hyperkeratosis; itching; non-infectious otitis externa; perianal inflammation; rash; scaling; skin blisters; skin irritation or burning; skin rash; swelling; weeping; worsening pruritic rash |
|  | Changes to Skin Barrier | Burn injuries; desquamation and wet-appearing skin with debris consisting of large amounts of gray to whitish desquamating skin; rectal burns; transient whitening of skin; whitish appearing excoriations |
|  | Features of Infection/Abscess | New onset anal ulcers; abscess-like presentation; phlegmonous changes; purulence |
| **Cardiovascular** | Cardiovascular | Cardiovascular symptoms; palpitations; erratic heart beats; cardiopulmonary; tachycardia |
| **Mortality** | Mortality | Death; early death; mortality |
| **Ocular** | Ocular | Blurred vision; eye irritation/burning; eye irritation, increased tearing |
| **Renal** | Renal | Acute renal failure; bladder infections; recurrent urinary tract infections; renal |
|  |  | disease; oliguric |
| **Miscellaneous** | Miscellaneous | Fever; febrile; methaemoglobinaemia |

*\*Unique health effects listed as stated in original studies*

**Table S4.** Quality Assessment of included Cohort & Case Control Studies.

| Author & Year | Q1 | Q2 | Q3 | Q4 | Q5 | Q6 | Q7 | Q8 | Q9 | Q10 | Q11 | Q12 | Q13 | Q14 | Q15 | Q16 | Q17 | Q18 | Q19 | Q20 | Q21 | Q22 | Score Absolute | Score (%) | Score Quality Category |
| --- | --- | --- | --- | --- | --- | --- | --- | --- | --- | --- | --- | --- | --- | --- | --- | --- | --- | --- | --- | --- | --- | --- | --- | --- | --- |
| Heaton et al., 2017 | 1 | 1 | 1 | 2 | 1 | 1 | 1 | 0 | 0 | 0 | 1 | 1 | 1 | 1 | 0 | 0 | 1 | 1 | 0 | 1 | 2 | 0 | 17 | 71 | Good |

**Table S5.**
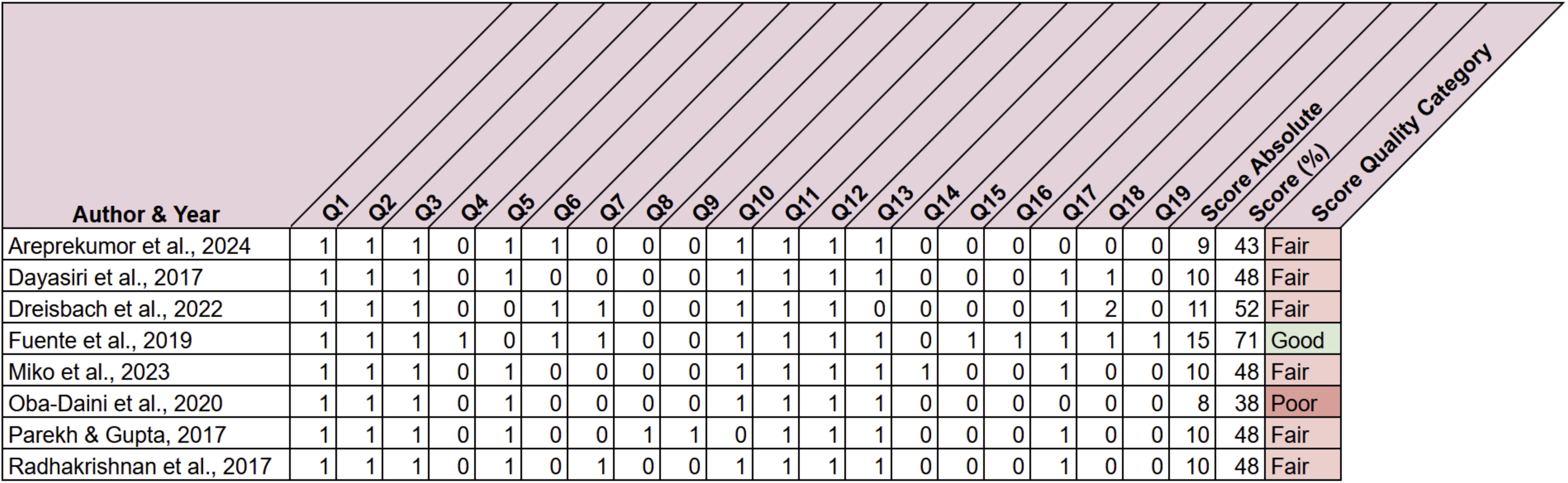
Quality Assessment of included Cross-Sectional Studies.

**Table S6.** Quality Assessment of included Case Reports & Case Studies Studies.

| Author & Year | Q1 | Q2 | Q3 | Q4 | Q5 | Q6 | Q7 | Score Absolute | Score (%) | Score Quality Category |
| --- | --- | --- | --- | --- | --- | --- | --- | --- | --- | --- |
| Benítez Riesco et al., 2024 | 1 | 2 | 1 | 1 | 1 | 0 | 1 | 7 | 70 | Good |
| Chattopadhyay et al., 2021 | 1 | 2 | 1 | 1 | 1 | 0 | 1 | 7 | 70 | Good |
| Contestable, 2017 | 1 | 2 | 1 | 1 | 1 | 1 | 1 | 8 | 80 | Good |
| Esashi et al., 2021 | 1 | 2 | 1 | 2 | 1 | 0 | 1 | 8 | 80 | Good |
| Fife et al., 2018 | 1 | 2 | 1 | 0 | 0 | 0 | 1 | 5 | 50 | Fair |
| Goenka et al., 2022 | 1 | 2 | 1 | 1 | 1 | 1 | 1 | 8 | 80 | Good |
| Gupta et al., 2017 | 1 | 2 | 1 | 1 | 1 | 0 | 1 | 7 | 70 | Good |
| Guss et al., 2020 | 1 | 2 | 1 | 1 | 1 | 0 | 1 | 7 | 70 | Good |
| Hara et al., 2018 | 1 | 2 | 1 | 1 | 1 | 1 | 1 | 8 | 80 | Good |
| Kim et al., 2018 | 2 | 2 | 1 | 2 | 1 | 0 | 1 | 9 | 90 | Excellent |
| Long & Charles, 2018 | 1 | 2 | 1 | 2 | 1 | 1 | 1 | 9 | 90 | Excellent |
| Oreh et al., 2023 | 1 | 2 | 1 | 2 | 1 | 0 | 1 | 8 | 80 | Good |
| Poon et al., 2019 | 1 | 2 | 1 | 2 | 1 | 0 | 1 | 8 | 80 | Good |
| Ravikanth et al., 2018 | 1 | 2 | 1 | 1 | 1 | 0 | 1 | 7 | 70 | Good |
| Salam et al., 2020 | 1 | 2 | 1 | 2 | 1 | 0 | 1 | 8 | 80 | Good |
| Sanju et al., 2020 | 2 | 2 | 1 | 2 | 1 | 0 | 1 | 9 | 90 | Excellent |

